# Kidney Function Estimation and its Relationship with Frailty in Older Adults: Insights from SHIP-TREND

**DOI:** 10.1101/2025.10.28.25339027

**Authors:** Yulia Komleva, Maximilian König, Till Ittermann, Nele Friedrich, Astrid Petersmann, Matthias Nauck, Henry Völzke, Marwan Mannaa, Maik Gollasch

## Abstract

**Background:** End-stage renal disease (ESRD) is closely associated with frailty, but the relationship between earlier stages of kidney dysfunction and frailty in the broader older population remains unclear. This study investigates the association between kidney function and frailty in community-dwelling older adults in Northeast Germany, using cross-sectional data from the population-based SHIP-TREND study. **Methods:** Data of 1,456 community-dwelling participants aged ≥60 years of the SHIP-TREND cohort were analyzed. Frailty was classified using modified Fried phenotype criteria. Kidney function was assessed using various estimated glomerular filtration rate (eGFR) formulas (MDRD, CKD-EPI and EKFC, based on creatinine or cystatin C). Associations between eGFR, and albuminuria and frailty were examined using multivariable linear and logistic regression models adjusted for age, sex, comorbidities, and body composition. **Results:** Frailty prevalence was 2.3%, and frail individuals had significantly lower eGFR, particularly when calculated using cystatin C-based formulas. CKD-EPI and EKFC equations using cystatin C showed the strongest associations with frailty (e.g. CKD-EPI_CysC_: OR = 31.3, 95 % CI 5.04 – 194.93, p < 0.001 for eGFR <30 vs. ≥60 mL/min/1.73 m²). Creatinine-based equations demonstrated weaker associations, likely due to confounding by muscle mass. Albuminuria was not significantly associated with frailty. Frail participants exhibited higher BMI but lower muscle mass, pointing to sarcopenic obesity as a potential contributor to the pathophysiology linking reduced kidney function and frailty. **Conclusions:** This study shows that lower kidney function, especially when estimated using cystatin C-based equations, is significantly associated with frailty in older adults. The strength of our study lies in the large, well-defined population and standardized assessments. In clinical practice, incorporating cystatin C into GFR estimation alongside muscle mass assessments may provide a more reliable framework for identifying CKD in older adults, and may also improve the prediction of frailty.

**Key points:**

- Lower kidney function, especially via cystatin C–based eGFR, is strongly linked to frailty in older adults.
- Sarcopenic obesity may drive this link, with frail individuals showing high BMI but low muscle mass.
- Combining cystatin C–based eGFR with muscle mass assessment may improve CKD detection and frailty prediction in clinical practice.

## INTRODUCTION

Chronic kidney disease (CKD) affects over 840 million people worldwide and is increasingly recognized not only as a driver of cardiovascular disease but also as a contributor to accelerated ageing and physical decline ^1^. End-stage renal disease (ESRD) is frequently accompanied by frailty ^1,2^, and adults of all ages undergoing hemodialysis have a high prevalence of frailty - more than five times as high as that observed among community-dwelling older adults ^3^. However, the link between earlier stages of kidney dysfunction and frailty in the general older population remains poorly understood ^2^.

In Germany, the prevalence of reduced kidney function is particularly high in the Northeast, with up to 25% of older adults (65–74 years) showing an estimated glomerular filtration rate (eGFR) <60 mL/min/1.73 m², which is substantially exceeding rates in other European populations^2^. However, eGFR estimation based on serum creatinine may be inaccurate in older individuals due to age-related changes in muscle mass ^4,5^. Cystatin C–based equations offer an alternative approach, potentially improving risk stratification in populations of older adults ^6,7^.

Given the considerable disease burden and regional discrepancies, further investigation is warranted to understand the underlying causes and implications of the elevated CKD prevalence observed in Northern Germany. Importantly, creatinine-based eGFR equations may underestimate renal impairment in older adults and individuals with sarcopenic obesity. ^4,5^. Therefore, incorporating alternative biomarkers or using other – novel - eGFR equations may provide a more accurate and nuanced assessment of kidney function across diverse subpopulations.

This study aimed to assess the prevalence of frailty and its association with key kidney function parameters, including eGFR, estimated using both creatinine- and cystatin C–based equations, as well as the urine albumin-creatinine ratio (UACR) in a representative population-based sample from Northeast Germany (SHIP-TREND cohort). We hypothesized that lower kidney function is significantly associated with frailty in older adults and that this relationship is modulated by body composition and the choice of biomarkers used to estimate GFR.

## METHODS

### Study population and design

The SHIP-TREND study (Study of Health in Pomerania – TREND) is a large-scale, population-based epidemiological cohort study conducted in Northeast Germany. As part of the broader SHIP project, SHIP-TREND was designed to assess the prevalence and incidence of common risk factors, subclinical disorders, and clinical diseases, as well as to investigate the complex associations among these health determinants ^8^. This cross-sectional analysis used data from the SHIP-TREND cohort, which has been described in detail previously ^9^. In total, baseline data were collected between 2008 and 2012 from 4,420 participants aged 20–79 years, randomly selected in an age- and sex-stratified manner from local residents’ registration offices in the cities of Stralsund, Greifswald, and Anklam and surrounding municipalities. For this cross-sectional analysis, we included community-dwelling participants aged over 60 years. After excluding individuals with missing information on frailty criteria, the final analytical sample comprised 1,456 participants. Of these, eGFR was available for 1,453 participants.

### Ethics statement

The study has been conducted according to the recommendations of the Declaration of Helsinki. The study protocol of SHIP-TREND was approved by the local ethics committee of the University of Greifswald (registration no. BB39/08) with all participants giving informed written consent.

### Key variables

#### Kidney function assessment

Serum creatinine and cystatin C were measured using the Dimension VISTA system (Siemens Healthcare Diagnostics, Eschborn, Germany). Creatinine was determined by a modified kinetic Jaffé method (measurement range: 0.14–20.2 mg/dL), and cystatin C by a nephelometric assay (limit of detection: 0.05 mg/L). Urinary creatinine and albumin concentrations were also measured using photometric and nephelometric assays, respectively, to calculate the urinary albumin-to-creatinine ratio (UACR; mg/g).

Estimated glomerular filtration rate (eGFR) was calculated using various equations: Modification of Diet in Renal Disease (MDRD), Chronic Kidney Disease Epidemiology Collaboration (CKD-EPI) creatinine- and cystatin C-based, and European Kidney Function Consortium (EKFC) creatinine- and cystatin C-based ^10–15^.

eGFR was categorized as <30, 30–44, 45–59, and ≥60 mL/min/1.73 m². CKD was defined as eGFR <60 mL/min/1.73 m². UACR was stratified into <30, 30–299, and ≥300 mg/g. For further details see the **Supplementary material**.

#### Frailty assessment

Frailty was operationalized according to the phenotype concept proposed by Fried et al. ^16^. Minor modifications to the original methodology were necessary to adjust for the data availability in SHIP-TREND. The five frailty phenotype criteria were assessed as follows:

◦ “Unintentional weight loss” was defined as a body mass index (BMI) of <18.5 kg/m², this criterion proved feasible and valid e.g. in the Atherosclerosis Risk in Communities (ARIC) Study ^17,18^.
◦ “Self-reported exhaustion”: Participants were classified as meeting the exhaustion criterion if they answered positively to at least one of the two relevant questions from the SF-12 questionnaire:

▪ *“ In the past four weeks, how often did you feel discouraged and sad?”*
▪ *“In the past four weeks, how often did you feel full of energy?”*
◦ “Weakness”: low handgrip strength (HGS) was measured by Smedley’s Dynamometer, Scandidact, Odder, Denmark ^16,19^. A criteria was defined using BMI- and sex-specific cut-off points, as described previously ^16^. For further details refer to the **Supplementary material.**
◦ “Slow walking speed”: walking speed was assessed in the Timed Up & Go test ^20^. A TUG time ≥19 seconds was used as the cut-off for criteria ^21^.
◦ “Low physical activity” was assessed based on self-reported participation in sports activities during the summer. Regular physical activity: ≥2 hours per week or 1–2 hours per week; low physical activity: <1 hour per week or no exercise.

Individuals were categorized as robust (0 criteria), prefrail (1–2 criteria), or frail (≥3 criteria). This classification method, adapted from the original frailty phenotype defined by Fried et al.^16^, provided an objective framework for identifying frailty in the SHIP-TREND cohort and examining its relationship with kidney function and other health parameters.

#### Covariables

Chronic diseases and socio-demographic variables were collected through computer-assisted personal interviews. Body composition parameters, including muscle mass and fat mass, were measured by multifrequency bioelectrical impedance analysis (Nutriguard M). BMI was calculated based on objectively measured height and weight, while waist circumference was determined using standard anthropometric techniques. Serum high-sensitivity C-reactive protein (hs-CRP) was measured using a immunonephelometric assay on the Dimension VISTA system (Siemens Healthcare Diagnostics, Eschborn, Germany) ^22^. For details see **Supplementary material (Methods).**

#### Statistical analysis

Median and interquartile range (Median [Q1; Q3]) were used to describe quantitative variables (eGFR, UACR, age). Frequencies and proportions (n [%]) were used to describe ordinal and categorical variables (Frailty status, eGFR group, UACR group).

Statistical tests, including the t-test, one-way analysis of variance (ANOVA), Kruskal‒Wallis test, Wilcoxon rank sum test, Fisher’s test, or χ2 test, were used as appropriate. Pearson correlation analysis was performed to assess the association between quantitative variables. Analysis of variance (ANOVA) with posterior pairwise comparisons was performed to compare three unrelated groups (frailty status) on quantitative variables (correction for multiple comparisons was performed using Bonferroni or Dunnett test).

Furthermore, regression analyses were performed: The primary exposures were eGFR and UACR. The primary outcome was frailty status (categorical; robust 0, pre-frail 1-2, frail ≥ 3), the secondary outcome was the frailty score (discrete; 0-5)

Univariable linear regression models were used to assess the association of eGFR with the discrete frailty score without adjustment for other variables. For adjusted analyses (adjustment for UACR and/or other variables (demographic domain: age, sex – Model 1; physical domain, comorbidity – Model 2; age, sex, comorbidity – Model 3), a multivariable linear regression models were used.

Logistic regression models were used to assess the association of eGFR and frailty (binary categorical variable: <3 or ≥3 points) a) without adjustment for other variables, and b) with adjustment for UACR and/or other variables (demographic domain: age, sex – Model 1; physical domain, comorbidity – Model 2; all variables – Model 3). Linear regression analyses were also applied to examine the association between eGFR, muscle mass, and their interaction in relation to frailty score.

At a significance level of p < 0.05, the null hypothesis was rejected, indicating a statistically significant result. Statistical analysis was performed using the STATA software package version (STATA/MP 18, StataCorp, Texas, USA).

## RESULTS

### Study population

A total of 1,456 participants were included in the analytical cohort, with eGFR estimated for 1,453 of them. **Table 1** summarizes the participants’ characteristics by frailty status (robust, pre-frail and frail). 33 participants (2.3%) were categorized as frail according to the Fried criteria, while 591 (40.7%) were pre-frail and 829 (57.0%) were robust (**Table 1**). The median age of robust participants was 68 years [64;72], and of frail participants 73 [68; 76] years. The prevalence of frailty was higher in women (3.5%), than in men (1.1%, p=0.003) (**Table 1**). Individuals who were classified as frail had slightly higher BMI 30.78 [28.68; 35.55] kg/m² than participants who were robust 28.62 [25.98; 31.44] kg/m² (p=0.026).

**Table 1.** — Cohort characteristics by frailty status.

|  | Robust | Pre-frail | Frail |
| --- | --- | --- | --- |
| n (%) | 829 (57.1%) | 591 (40.7%) | 33 (2.3%) |
| Age, years | 68.00 [64.00; 72.00] | 69.00 [64.00; 75.00] | 73.00 [68.00; 76.00] |
| Female | 399 (55.7%) | 292 (40.8%) | 25 (3.5%) |
| Male | 431 (58.4%) | 299 (40.5%) | 8 (1.1%) |
| BMI, kg/m<sup>2</sup> | 28.61 [25.98; 31.44] | 29.62 [26.82; 33.05] | 30.78 [28.68; 35.55] |
| eGFR MDRD<sub>SCr</sub> | 75.60 [64,50; 87,90] | 72.70 [62,20; 84,70] | 74.60 [57,50; 79,10] |
| eGFR CKD-EPI<sub>SCr</sub> | 78.69 [66.15; 91.10] | 75.33 [63.33; 88.31] | 76.88 [59.76; 81.60] |
| eGFR CKD-EPI<sub>CysC</sub> | 98.70 [84.70; 106.57] | 93.62 [76.77; 103.90] | 84.66 [74.20; 98.20] |
| eGFR EKFC<sub>SCr</sub> | 69.03 [57.88; 79.28] | 66.58 [55.44; 76.23] | 65.52 [52.99; 70.81] |
| eGFR EKFC<sub>CysC</sub> | 84.76 [78.71; 89.76] | 82.27 [74.47; 88.65] | 78.89 [73.55; 84.76] |
| UACR, mg/mmol | 1.43 [0.79; 3.19] | 1.51 [0.80; 3.22] | 1.67 [1.04; 3.72] |
| UACR, mg/g | 12.62 [6.96; 28.17] | 13.32 [7.09; 28.46] | 14.73 [9.18; 32.92] |
| C-reactive protein, mg/L | 1.43 [0.78; 2.73] | 1.77 [0.91; 3.66] | 1.89 [1.13; 3.14] |
| Muscle weight, kg | 56.00 [47.10; 65.60] | 54.50 [46.50; 65.00] | 49.30 [44.80; 56.30] |
| Number of diseases<br>Comorbidities | 2 [1; 3] | 2 [1; 3] | 3 [2; 4] |

Notes and abbreviations: Baseline demographic, clinical, and laboratory characteristics of participants across frailty categories. Median [Q1; Q3] values are shown for continuous variables.
Numbers and percentages are shown for categorical variables. *eGFR values are expressed in mL/min/1.73 m<sup>2</sup>. BMI: body mass index; eGFR: estimated glomerular filtration rate; MDRD: Modification of Diet in Renal Disease equation; CKD-EPI: Chronic Kidney Disease Epidemiology Collaboration equation; EKFC: European Kidney Function Consortium equation; eGFR<sub>cr</sub>: creatinine-based eGFR; eGFR<sub>cys</sub>: cystatin C-based eGFR; UACR: urinary albumin-to-creatinine ratio.*

### Kidney function

A significantly reduced eGFR among frail participants was particularly evident with cystatin C-based eGFR (eGFR_CysC_), compared to creatinine-based estimates (eGFR_SCr_) using the MDRD or CKD-EPI equations, where eGFR remained relatively stable across all frailty categories. E.g. the mean eGFR_CysC_ according to CKD-EPI in frail participants was 84.66 [74.20; 98.20] vs. 98.70 [84.70; 106.57] ml/min/1.73m^2^ in robust individuals. The cystatin C-based EKFC equation provided a similar pattern: 84.76 [78.71; 89.76] in robust individuals vs. 78.89 [73.55; 84.76] ml/min/1.73m^2^ in frail individuals, whereas the difference was much smaller with creatinine-based MDRD or CKD-EPI formula (**Table 1**).

### Co-factors

On average, the C-reactive protein increased stepwise from 1.43 [0.78; 2.73] mg/L in robust individuals to 1.89 [1.13; 3.14] mg/L in frail patients. Frail individuals showed significantly lower muscle mass, being 49.3 [44.80; 56.30] kg, compared to 56.0 [47.10; 65.60] kg in robust participants (p=0.026).

Frail individuals tended to have a higher BMI (30.78 [28.68; 35.55] kg vs. 28.62 [25.98; 31.44] kg in robust and 29.65 [26.85; 33.05] kg in pre-frail participants (p<0.001)) (**Table 1**). In addition, the relation of muscle mass relative to total body weight declined progressively with increasing frailty severity (p < 0.001). A post hoc analysis is presented in **Supplementary Table 1**. To further quantify this association, correlation analyses were conducted with the frailty score, demonstrating a negative correlation between frailty severity and muscle mass (r = −0.073, p = 0.007) (**Supplementary Table 2)**. Also, the median comorbidity count was higher in frail participants than in the other groups reflecting the increased burden of multimorbidity in frailty (**Table 1**).

In linear regression models with the frailty score as the dependent variable, and eGFR, muscle mass, and their interaction term (eGFR × muscle mass) as exposures, across all models, lower eGFR was significantly associated with higher frailty scores, with β ranging from –0.011 (MDRD_SCr_) to –0.030 (EKFC_CysC_). Muscle mass also showed a significant inverse association with frailty in most models **(Supplementary Table 3**). Only in the EKFC_CysC_ model, did the interaction term reach significance (p = 0.046), indicating that the relationship between kidney function and frailty varied with muscle mass.

While the inverse association between the frailty score and eGFR was consistently observed irrespective of the formula used for estimation (all *p* < 0.001, **Table 2**), however, the strength of the association varied across formulas, with the EKFC_CysC_ formula exhibiting the strongest association with frailty (ß = −0.012 [−0.016; −0.009], p < 0.001).

**Table 2.**
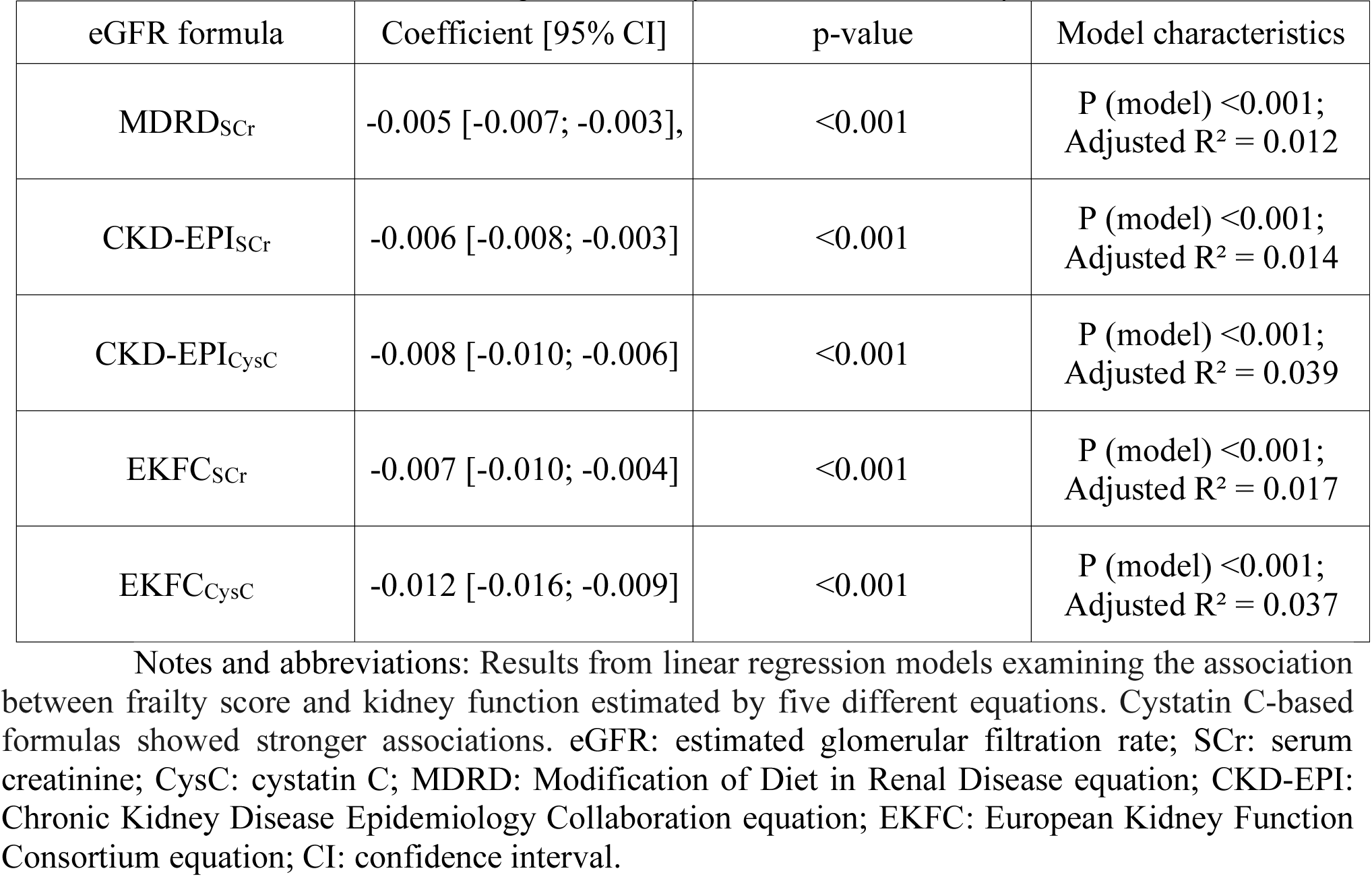
— Univariate linear regression analysis of eGFR and frailty score.

### Multivariable analyses of the association between kidney function and frailty

In multivariable analyses, adjusted for confounding factors, eGFR equations based on creatinine remained significantly associated with frailty (**Table 3**). Lower eGFR_SCr_, calculated using the MDRD, CKD-EPI, and EKFC formulas—were consistently linked to higher frailty scores in models adjusted for demographic factors (Model 1) and comorbidity (Model 2), regardless of UACR inclusion. However, the strength of these associations diminished for creatinine-based formulas after adjustment for all covariates (Model 3).

**Table 3.** — Multivariable linear regression analysis of eGFR and frailty score.

|  | Coefficient [95% CI], P-value | Model characteristics |
| --- | --- | --- |
| **eGFR MDRD<sub>SCr</sub>** |  |  |
| *Multivariable linear regression: models without UACR* |  |  |
| eGFR adjusted for sex, age (Model 1) | -0.003 [-0.005; -0.001],<br>P = 0.011 | P-value (model) <0.001<br>Adjusted R<sup>2</sup> = 0.027 |
| eGFR adjusted for comorbidity (Model 2) | -0.003 [-0.006; -0.001],<br>P = 0.001 | P-value (model) <0.001<br>Adjusted R<sup>2</sup> = 0.026 |
| eGFR adjusted for sex, age, comorbidity (Model 3) | -0.002 [-0.004; 0.000],<br>P = 0.064 | P-value (model) <0.001<br>Adjusted R<sup>2</sup> = 0.037 |
| *Multivariable linear regression: models with UACR* |  |  |
| eGFR adjusted for UACR, sex, age | -0.003 [-0.005; -0.001],<br>P = 0.033 | P-value (model) <0.001<br>Adjusted R<sup>2</sup> = 0.028 |
| eGFR adjusted for UACR, comorbidity | -0.004 [-0.006; -0.001],<br>P = 0.002 | P-value (model) <0.001<br>Adjusted R<sup>2</sup> = 0.019 |
| eGFR adjusted for UACR, sex, age, comorbidity | -0.002 [-0.005; 0.000],<br>P = 0.101 | P-value (model) <0.001<br>Adjusted R<sup>2</sup> = 0.032 |
| **eGFR (CKD-EPI<sub>SCr</sub>)** |  |  |
| *Multivariable linear regression: models without UACR* |  |  |
| eGFR adjusted for sex, age | -0.003 [-0.006; -0.001]; p = 0.018 | P (model) <0.001; Adjusted R2 = 0.027 |
| eGFR adjusted for comorbidity | -0.004 [-0.007; -0.002]; p <0.001 | P (model) <0.001; Adjusted R2 = 0.027 |
| eGFR adjusted for sex, age, comorbidity | -0.002 [-0.005; 0.000]; p = 0.093 | P (model) <0.001; Adjusted R2 = 0.037 |
| *Multivariable linear regression: models with UACR (mg/g)* |  |  |
| eGFR adjusted for UACR, sex, age | -0.003 [-0.006; 0.000]; p = 0.034 | P (model) <0.001; Adjusted R2 = 0.028 |
| eGFR adjusted for UACR, comorbidity | -0.005 [-0.008; -0.002]; p = 0.001 | P (model) <0.001; Adjusted R2 = 0.021 |
| eGFR adjusted for UACR, sex, age, comorbidity | -0.002 [-0.006; 0.001]; p = 0.108 | P (model) <0.001; Adjusted R2 = 0.032 |
| **eGFR (CKD-EPI<sub>CysC</sub>)** |  |  |
| *Multivariable linear regression: models without UACR* |  |  |
| eGFR adjusted for sex, age | -0.007 [-0.009; -0.004]; p <0.001 | P (model) <0.001; Adjusted R2 = 0.043 |
| eGFR adjusted for comorbidity | -0.007 [-0.009; -0.005]; p <0.001 | P (model) <0.001; Adjusted R2 = 0.047 |
| eGFR adjusted for sex, age, comorbidity | -0.006 [-0.008; -0.003]; p <0.001 | P (model) <0.001; Adjusted R2 = 0.050 |
| *Multivariable linear regression: models with UACR (mg/g)* |  |  |
| eGFR adjusted for UACR, sex, age | -0.006 [-0.009; -0.004]; p <0.001 | P (model) <0.001; Adjusted R2 = 0.041 |
| eGFR adjusted for UACR, comorbidity | -0.007 [-0.010; -0.005]; p <0.001 | P (model) <0.001; Adjusted R2 = 0.040 |
| eGFR adjusted for UACR, sex, age, comorbidity | -0.006 [-0.009; -0.003]; p <0.001 | P (model) <0.001; Adjusted R2 = 0.044 |
| **eGFR (EKFC<sub>Scr</sub>)** |  |  |
| *Multivariable linear regression: models without UACR* |  |  |
| eGFR adjusted for sex, age | -0.004 [-0.007; -0.001]; p = 0.018 | P (model) <0.001; Adjusted R2 = 0.027 |
| eGFR adjusted for comorbidity | -0.005 [-0.008; -0.003]; p <0.001 | P (model) <0.001; Adjusted R2 = 0.029 |
| eGFR adjusted for sex, age, comorbidity | -0.003 [-0.006; 0.000]; p = 0.094 | P (model) <0.001; Adjusted R2 = 0.037 |
| *Multivariable linear regression: models with UACR (mg/g)* |  |  |
| eGFR adjusted for UACR, sex, age | -0.004 [-0.007; 0.000]; p = 0.036 | P (model) <0.001; Adjusted R2 = 0.028 |
| eGFR adjusted for UACR, comorbidity | -0.006 [-0.009; -0.003]; p <0.001 | P (model) <0.001; Adjusted R2 = 0.023 |
| eGFR adjusted for UACR, sex, age, comorbidity | -0.003 [-0.007; 0.001]; p = 0.113 | P (model) <0.001; Adjusted R2 = 0.032 |
| **eGFR (EKFC<sub>CysC</sub>)** |  |  |
| *Multivariable linear regression: models without UACR* |  |  |
| eGFR adjusted for sex, age | -0.012 [-0.016; -0.008]; p <0.001 | P (model) <0.001; Adjusted R2 = 0.043 |
| eGFR adjusted for comorbidity | -0.011 [-0.014; -0.007]; p <0.001 | P (model) <0.001; Adjusted R<sup>2</sup> = 0.045 |
| eGFR adjusted for sex, age, comorbidity | -0.010 [-0.015; -0.006]; p <0.001 | P (model) <0.001; Adjusted R<sup>2</sup> = 0.049 |
| *Multivariable linear regression: models with UACR (mg/g)* |  |  |
| eGFR adjusted for UACR, sex, age | -0.011 [-0.016; -0.006]; p <0.001 | P (model) <0.001; Adjusted R<sup>2</sup> = 0.042 |
| eGFR adjusted for UACR, comorbidity | -0.011 [-0.015; -0.007]; p <0.001 | P (model) <0.001; Adjusted R<sup>2</sup> = 0.038 |
| eGFR adjusted for UACR, sex, age, comorbidity | -0.010 [-0.015; -0.005]; p <0.001 | P (model) <0.001; Adjusted R<sup>2</sup> = 0.044 |

**Notes and abbreviations:** Multivariable models adjusted for age, sex, comorbidities, and UACR. The strength of association between eGFR and frailty persisted predominantly for cystatin C-based formulas. Coefficients represent the estimated change in frailty score per 1 mL/min/1.73 m<sup>2</sup> increase in eGFR. Each formula was analyzed in separate models adjusted for various covariates, with and without inclusion of albuminuria (UACR). *eGFR: estimated glomerular filtration rate; SCr: serum creatinine; CysC: cystatin C; MDRD: Modification of Diet in Renal Disease; CKD-EPI: Chronic Kidney Disease Epidemiology Collaboration; EKFC: European Kidney Function Consortium; UACR: urine albumin-to-creatinine ratio (mg/g); CI: confidence interval.*

Overall, cystatin C-based equations (CKD-EPI and EKFC) showed stronger associations between kidney function and frailty, even after adjustment for age, sex, comorbidities (Models 1, 2, 3), and UACR. Among these, the EKFC_CysC_ equation emerged as having the strongest association with frailty (**Table 3**).

### Frailty and CKD stages

Severely decreased eGFR (<30 ml/min/1.73 m²) was common among frail individuals. The proportion with eGFR below this threshold varied depending on the equation used: 18.2% for EKFC_SCr_, 25% for MDRD_SCr_ and CKD-EPI_SCr,_ 33.3% for EKFC_CysC_, and 40% for CKD-EPI_SCysC_ (**Supplementary, Table 4).**

When classified into four groups based on kidney function (*normal and mildly decreased, G1-G2: eGFR ≥ 60 mL/min/1.73 m*^2^*; mild-moderate decreased: eGFR 45–59 mL/min/1.73 m*^2^ *, G3a; moderate-severe decrease: eGFR 30–44 mL/min/1.73 m*^2^*, G3b; severely decreased: eGFR <30 mL/min/1.73 m*^2^*, G4-G5*) the proportion with eGFR≥60 mL/min/1.73m² (G1-G2) was highest when estimated using EKFC_CysC_ (72.7%) and CKD-EPI_CysC_ (71.8%). Also, when classified according to the KDIGO system based on eGFR and albuminuria, CKD-EPI_CysC_ and EKFC_CysC_ equations resulted in consistently lower proportions of participants in advanced CKD stages (≥A2 and ≥G3b) – potentially reflecting more conservative estimates in populations of older adults (**Supplementary Table 5**). There was also evidence of a dose–response relationship between CKD severity and frailty prevalence. Frailty prevalence ranged from 2.0–2.1% among those with GFR ≥ 60, 2.6–3.8% for GFR 45–59, 0–5.3% for GFR 30–44, to 18.2–40% for GFR < 30 mL/min/1.73m², depending on the equation used. (**Supplementary Table 4**).

For each eGFR formula, the unadjusted prevalence of frailty markedly increased as the eGFR decreased below 30 mL/min/1.73m². The strongest association with frailty was observed using CKD-EPI_CysC_ (eGFR < 30 vs. eGFR >= 60 OR = 31.3, 95 % CI 5.04 – 194.93, p < 0.001), indicating that individuals with eGFR <30 mL/min/1.73m² were considerably more likely to be frail compared with those with preserved renal function. The EKFC_CysC_ formula also showed a strong association (OR = 23.0, 95 % CI 2.03 – 260.86, p = 0.011), though slightly lower than for CKD-EPI_CysC_. For eGFR values between 30–44 and 45–59 mL/min/1.73m², none of the formulas showed significant associations with frailty, indicating that the link between kidney function and frailty is particularly pronounced with severe CKD (eGFR <30 mL/min/1.73m²) (**Table 4**).

**Table 4.**
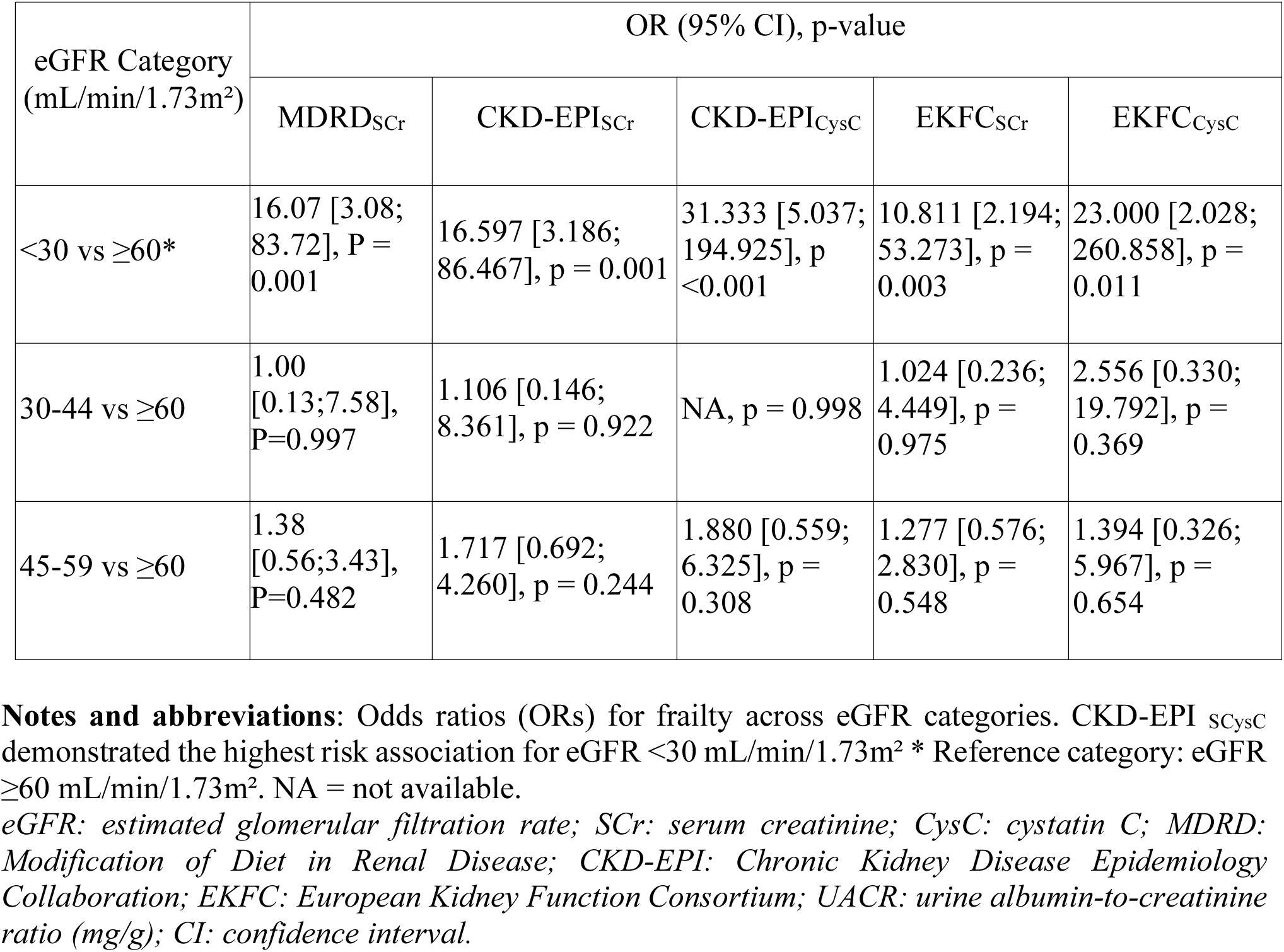
— Univariable logistic regression analysis of eGFR and frailty (frailty score ≥3)

Likewise, in multivariable logistic regression analyses an eGFR_SCr_ (MDRD and CKD-EPI) of <30 ml/min/1.73m^2^ was significantly associated with frailty in all models (**Supplementary Tables 6a-d**). Similarly, when kidney function was estimated using EKFC_SCr_, an eGFR <30 ml/min/1.73 m² was significantly associated with frailty only in Model 2 (adjusted for comorbidity), but not in the fully adjusted model (**Supplementary Table 6g-h**).

When kidney function was estimated using CKD-EPI_CysC_ or EKFC_CysC_, eGFR <30 ml/min/1.73 m² remained significantly associated with frailty in Model 1, and this association persisted in Model 3, with and without UACR adjustment, reinforcing the robustness of the relationship between severe renal impairment and frailty (**Supplementary Table 6e-f**). Overall, CKD-EPI_CysC_ demonstrated the strongest association between (low) eGFR and frailty, with odds ratios exceeding 20 in some models (**Supplementary Table 6i-j**). Interestingly, across all formulas, female sex and older age were strong predictors, with women exhibiting approximately a fourfold increase in the odds of frailty.

Interestingly, both moderate (ACR 30–299 mg/g) and severe (ACR ≥300 mg/g) albuminuria did not exhibit a significant association with frailty (**Supplementary Table 6a-j)**. This is consistent with the finding, that overall, adjusting for UACR had minimal impact on the associations, reinforcing that eGFR is an independent predictor of frailty.

## DISCUSSION

### Prevalence and renal correlates of frailty in the SHIP-TREND study population

In this population-based cohort of older adults (SHIP-TREND), we observed that only 2.3% of older adults between 64 and 84 years were frail according to Fried’s phenotype criteria, while 40.7% were classified as pre-frail. Frailty was significantly more common among women and increased with age, which is consistent with previous research findings ^23^. On average, frail individuals exhibited a distinct phenotypic profile characterized by elevated BMI alongside markedly reduced absolute and relative muscle mass. This supports the concept of sarcopenic obesity as a central feature of frailty^24^ and together with the age- and sex-distribution reinforces the validity of our frailty phenotype, providing confidence in its use for subsequent analyses of frailty’s associations with renal function ^25^.

Lower eGFR was consistently associated with greater frailty. The proportion of frail participants was higher among those with eGFR 45–59, 30–44, and <30 mL/min/1.73 m² compared to those with eGFR ≥ 60 mL/min/1.73m², although a statistically significant association was only observed for eGFR <30 mL/min/1.73 m². In individuals with severe kidney impairment (eGFR <30 mL/min/1.73 m²), the proportion of participants with frailty was particularly high - ranging from 18.2% to 40%, depending on the equation applied. Frailty was considerably less common in those with moderate CKD (eGFR 30–59 mL/min/1.73 m²) and was lowest among individuals with preserved renal function (eGFR ≥60 mL/min/1.73 m²). This dose–response pattern is consistent with prior studies demonstrating an increasing prevalence of frailty with advancing CKD severity^6,26–28^. While our study demonstrates a consistent inverse association between eGFR and frailty, regardless of the equation used to estimate kidney function. However, the magnitude of this association varied. Specifically, cystatin C–based estimations (CKD-EPI and EKFC) showed stronger relationships with frailty compared to creatinine-based GFR estimates. This finding aligns with earlier reports that in older adults, especially those with sarcopenia, creatinine-based GFR equations can substantially overestimate kidney function ^10,11,29–33^.

Although cystatin C is generally viewed as largely independent of muscle mass, its levels can still be affected by non-GFR factors such as inflammation, smoking, adiposity, and thyroid function ^11,31,34,35^. Particularly, in older or chronically ill individuals, complex physiological interactions may influence its validity as a marker of kidney function. Further, low muscle mass and reduced kidney function may co-occur, not due to a direct link between cystatin C and muscle mass, but because both are influenced by shared underlying processes. Indeed, in our cross-sectional analysis, cystatin C– based eGFR showed a significant association with muscle mass. The marginal interaction observed suggests that the relationship between reduced kidney function and frailty may differ by muscle mass level. This underscores the need to consider both renal biomarkers and body composition in geriatric risk assessment ^36–38^.

While continuous improvements in GFR estimation equations are valuable, it is essential to recognize that these equations provide only an approximation of the true renal function ^6^. In line with our findings, eGFR calculations based on cystatin C demonstrated a stronger and more consistent association with frailty than creatinine-based estimates, underlining the potential clinical advantage of cystatin C in assessing renal function among older adults. Nevertheless, direct measurement of GFR using exogenous filtration markers remains indispensable for precise evaluation of kidney function. Future studies should aim to refine both estimated and measured GFR approaches, promote broader application of iohexol clearance, and advance the development of rapid and reliable bedside assessment tools. Ultimately, measured and estimated GFR should be viewed as complementary rather than competing strategies in the comprehensive assessment of renal function ^39^. Interestingly, despite the strong link between kidney function and frailty, we found no significant association between albuminuria and frailty - possibly due to low case numbers limiting interpretability. While prior studies suggest albuminuria may contribute to frailty ^28,40,41^, further research, particularly longitudinal and mechanistic studies, is needed to clarify this potential link.

### Limitations of BMI and the need for muscle-sensitive renal assessment in older adults

Our findings also reinforce the complex interplay between GFR, body composition and frailty. While frail individuals exhibited a higher BMI, they also demonstrated significantly lower muscle mass. This highlights the limitations of BMI as a sole indicator of body composition – e.g. for adjusting GFR - and underscores the importance of assessing muscle mass directly using bioelectrical impedance analysis (BIA) or other objective measures, such as dual x-ray absorptiometry. The observed inverse association between frailty and muscle mass, coupled with a positive association between high BMI and frailty, supports the hypothesis that sarcopenic obesity plays a crucial role in frailty development ^42^. While BMI ≥25 kg/m² is generally associated with a 43% increased risk of frailty ^43^, our data support a U-shaped relationship, where both high and low BMI values are linked to frailty.

This is especially important for kidney function assessment. Sarcopenia can lead to overestimation of GFR with creatinine-based equations, masking early impairment ^44^. Indeed, although, cystatin C-based eGFR is considered less dependent on muscle mass, also correlated with muscle mass – likely due to indirect links with inflammation and obesity ^45^. In older adults, where muscle loss often occurs alongside systemic changes, alterations in body composition may distort renal function estimates, affecting diagnosis and therapeutic management. Both, from a clinical and public health perspective, using cystatin C - alone or with creatinine in race-free equations - may improve risk detection and care in older adults by offering a more accurate assessment of kidney function ^14^. However, this assumption needs validation against directly measured GFR using exogenous markers.

### Strengths and limitations

This study benefits from the robust, population-based design of the SHIP-TREND cohort with comprehensive phenotyping and objective muscle mass measurement, which improves confidence in both frailty classification and renal function estimation. The parallel use of creatinine- and cystatin C–based eGFR estimates provides complementary insights into kidney function, particularly in the context of age-related muscle loss, and highlights the differences between these biomarkers. However, the cross-sectional design limits causal interpretation, and the relatively low prevalence of frailty may have reduced statistical power and limited the feasibility of detailed subgroup analyses. Moreover, cystatin C levels can be affected by non-renal factors, and our frailty assessment focused mainly on physical aspects, potentially overlooking cognitive and social dimensions that may also interact with kidney function. Furthermore, one of the frailty criteria applied in this study was a BMI below 18.5 kg/m². At the same time, many frail individuals in our cohort exhibited elevated BMI values. This discrepancy may have influenced the observed distribution and limited our ability to fully capture the characteristic U-shaped relationship between body weight and frailty. If unintentional weight loss (e.g., ≥5% over recent months) had been available as a frailty indicator, it is likely that a larger number of participants would have met frailty criteria, potentially strengthening the observed association and improving the detection of both ends of the BMI spectrum related to frailty risk.

### Conclusions

This study shows that a reduced glomerular filtration rate - particularly when estimated using cystatin C – is consistently associated with frailty in older adults. In frail individuals, decreased kidney function may contribute to frailty through mechanisms such as protein-energy wasting, chronic inflammation, and hormonal changes, all of which are also reflected in altered body composition. Accordingly, frailty should be acknowledged as an additional major adverse aspect of CKD, together with cardiovascular disease and elevated mortality risk. In clinical practice, incorporating cystatin C-based GFR estimations alongside muscle mass assessments may provide a more reliable framework for identifying CKD in older adults, and may also improve the prediction of frailty.

## Data Availability

All data produced in the present study are available upon reasonable request to the authors

## Acknowledgements

The authors express their sincere gratitude to Irina Berdalina for her valuable consulting assistance with the statistical data analysis. During the preparation of this manuscript, the authors used ChatGPT4 in order to improve language and readability. After using the tool/service, the authors reviewed and edited the content as needed and take full responsibility for the content of the published article.

## Author Contributions

Conceptualization: Yulia Komleva, Maximilian König, Maik Gollasch

Data curation: Maximilian König, Till Ittermann, Nele Friedrich

Formal analysis: Yulia Komleva, Maximilian König, Till Ittermann

Methodology: Maximilian König, Till Ittermann, Astrid Petersmann

Supervision: Maximilian König, Henry Völzke, Matthias Nauck

Funding acquisition: Maik Gollasch

Investigation: Yulia Komleva

Project administration: Maik Gollasch

Writing – original draft: Yulia Komleva, Maximilian König

Writing – review & editing: Yulia Komleva, Maximilian König, Till Ittermann, Nele Friedrich, Astrid Petersmann, Matthias Nauck, Henry Völzke, Marwan Mannaa, Maik Gollasch

## Data Sharing Statement

Partial restrictions to the data and/or materials apply. We are not permitted to share individual data from the current study, but we are open to collaborative research with researchers worldwide, who can have access to analyzed data from our university.

## Supplementary material (Methods)

Kidney function assessment, C-reactive protein, Socio-demographic variables and body composition, Handgrip strength

## Supplementary tables

**Table 1.** Body composition parameters across frailty phenotypes

**Table 2.** Correlation between frailty score and anthropometric measures

**Table 3.** Linear regression models assessing the association between frailty score, eGFR, muscle mass, and their interaction

**Table 4.** Participant characteristics stratified by frailty and CKD stage

**Table 5.** Prevalence of CKD categories based on eGFR and UACR

**Figure 1.** Frailty prevalence across eGFR categories based on different estimation equations

**Table 6.** Multivariable associations between kidney function and frailty

**Table 6 a.** Association between eGFR MDRD _SCr_ and frailty (frailty score ≥3) adjusted for selected confounders

**Table 6 b.** Associations between measures of kidney function eGFR CKD-EPI_SCr_ and frailty score ≥3 and other confounders: models with UACR

**Table 6 c.** Associations between measures of kidney function eGFR CKD-EPI_SCr_ and frailty score ≥3 and other confounders: models without UACR

**Table 6 d.** Associations between measures of kidney function eGFR CKD-EPI_SCr_ and frailty score ≥3 and other confounders: models with UACR

**Table 6 e.** Associations between measures of kidney function eGFR CKD-EPI_CysC_ and frailty score ≥3 and other confounders: models without UACR

**Table 6 f.** Associations between measures of kidney function eGFR CKD-EPI_CysC_ and frailty score ≥3 and other confounders: models with UACR

**Table 6 g.** Associations between measures of kidney function eGFR EKFC_SCr_ and frailty score ≥3 and other confounders: models without UACR

**Table 6 h.** Associations between measures of kidney function eGFR EKFC_SCr_ and frailty score ≥3 and other confounders: models with UACR

**Table 6 i.** Associations between measures of kidney function eGFR EKFC_CysC_ and frailty score ≥3 and other confounders: models without UACR

**Table 6 j.** Associations between measures of kidney function eGFR EKFC_CysC_ and frailty score ≥3 and other confounders: models with UACR

---

**Figure 1.** Frailty prevalence across eGFR categories based on different estimation equation. The figure illustrates the proportion of frail individuals across various eGFR categories (G1–G5) as estimated by five equations: MDRD_SCr_, CKD-EPI_SCr_, CKD-EPI_CysC_, EKFC_SCr,_ and EKFC_CysC_. A consistent trend of higher frailty prevalence with declining eGFR was observed across all methods, with the CKD-EPI _SCysC_ and EKFC _SCysC_ equations identifying the highest proportions. eGFR: estimated glomerular filtration rate; SCr: serum creatinine; CysC: cystatin C; MDRD: Modification of Diet in Renal Disease; CKD-EPI: Chronic Kidney Disease Epidemiology Collaboration; EKFC: European Kidney Function Consortium.

## Reference

1. Kovesdy CP. Epidemiology of chronic kidney disease: an update 2022. Kidney Int Suppl. 2022;12(1):7–11. doi:10.1016/j.kisu.2021.11.003

2. Duarte MP, Almeida LS, Neri SGR, et al. Prevalence of sarcopenia in patients with chronic kidney disease: a global systematic review and meta-analysis. J Cachexia Sarcopenia Muscle. 2024;15(2):501–512. doi:10.1002/jcsm.13425

3. McAdams-DeMarco MA, Law A, Salter ML, et al. Frailty as a Novel Predictor of Mortality and Hospitalization in Individuals of All Ages Undergoing Hemodialysis. J Am Geriatr Soc. 2013;61(6):896–901. doi:10.1111/jgs.12266

4. Patel SS, Molnar MZ, Tayek JA, et al. Serum creatinine as a marker of muscle mass in chronic kidney disease: results of a cross-sectional study and review of literature. J Cachexia Sarcopenia Muscle. 2013;4(1):19–29. doi:10.1007/s13539-012-0079-1

5. Nankivell BJ, Nankivell LFJ, Elder GJ, Gruenewald SM. How unmeasured muscle mass affects estimated GFR and diagnostic inaccuracy. EClinicalMedicine. 2020;29–30:100662. doi:10.1016/j.eclinm.2020.100662

6. Dalrymple LS, Katz R, Rifkin DE, et al. Kidney Function and Prevalent and Incident Frailty. Clin J Am Soc Nephrol. 2013;8(12):2091–2099. doi:10.2215/CJN.02870313

7. Hart A, Blackwell TL, Paudel ML, et al. Cystatin C and the Risk of Frailty and Mortality in Older Men. J Gerontol A Biol Sci Med Sci. Published online November 9, 2016:glw223. doi:10.1093/gerona/glw223

8. Lüdtke L, Ittermann T, Großjohann R, et al. Risk Factors of Age-Related Macular Degeneration in a Population-Based Study: Results from SHIP-TREND-1 (Study of Health in Pomerania-TREND-1). Med Sci Monit. 2024;30. doi:10.12659/MSM.943140

9. Völzke H, Schössow J, Schmidt CO, et al. Cohort Profile Update: The Study of Health in Pomerania (SHIP). Int J Epidemiol. 2022;51(6):e372–e383. doi:10.1093/ije/dyac034

10. Pottel H, Björk J, Rule AD, et al. Cystatin C–Based Equation to Estimate GFR without the Inclusion of Race and Sex. N Engl J Med. 2023;388(4):333–343. doi:10.1056/NEJMoa2203769

11. Pottel H, Delanaye P, Cavalier E. Exploring Renal Function Assessment: Creatinine, Cystatin C, and Estimated Glomerular Filtration Rate Focused on the European Kidney Function Consortium Equation. Ann Lab Med. 2024;44(2):135–143. doi:10.3343/alm.2023.0237

12. Hennings A, Hannemann A, Rettig R, et al. Circulating Angiopoietin-2 and Its Soluble Receptor Tie-2 Concentrations Are Related to Renal Function in Two Population-Based Cohorts. Shimosawa T, ed. PLOS ONE. 2016;11(11):e0166492. doi:10.1371/journal.pone.0166492

13. Levey AS, Coresh J, Balk E, et al. National Kidney Foundation Practice Guidelines for Chronic Kidney Disease: Evaluation, Classification, and Stratification. Ann Intern Med. 2003;139(2):137–147. doi:10.7326/0003-4819-139-2-200307150-00013

14. Inker LA, Eneanya ND, Coresh J, et al. New Creatinine- and Cystatin C–Based Equations to Estimate GFR without Race. N Engl J Med. 2021;385(19):1737–1749. doi:10.1056/NEJMoa2102953

15. Inker LA, Schmid CH, Tighiouart H, et al. Estimating Glomerular Filtration Rate from Serum Creatinine and Cystatin C. N Engl J Med. 2012;367(1):20–29. doi:10.1056/NEJMoa1114248

16. Fried LP, Tangen CM, Walston J, et al. Frailty in Older Adults: Evidence for a Phenotype. J Gerontol A Biol Sci Med Sci. 2001;56(3):M146–M157. doi:10.1093/gerona/56.3.M146

17. Rutherford M, Downer B, Li CY, Chou LN, Al Snih S. Body mass index and physical frailty among older Mexican Americans: Findings from an 18-year follow up. Abete P, ed. PLOS ONE. 2022;17(9):e0274290. doi:10.1371/journal.pone.0274290

18. Huxley RR, Misialek JR, Agarwal SK, et al. Physical Activity, Obesity, Weight Change, and Risk of Atrial Fibrillation: The Atherosclerosis Risk in Communities Study. Circ Arrhythm Electrophysiol. 2014;7(4):620–625. doi:10.1161/CIRCEP.113.001244

19. Schulte S, Ittermann T, Gross S, et al. The relationship between age related changes in strength and fitness with body size, shape and composition. Sci Rep. 2025;15(1):9833. doi:10.1038/s41598-025-93828-2

20. Podsiadlo D, Richardson S. The Timed “Up & Go”: A Test of Basic Functional Mobility for Frail Elderly Persons. J Am Geriatr Soc. 1991;39(2):142–148. doi:10.1111/j.1532-5415.1991.tb01616.x

21. König M, Malsch C, Mariño J, et al. Nocturia as a clinical marker of loss of function and resilience or risk factor for frailty in older adults? Results of the Berlin Aging Study II. GeroScience. Published online January 31, 2025. doi:10.1007/s11357-025-01525-9

22. Braisch U, Koenig W, Rothenbacher D, et al. N-terminal pro brain natriuretic peptide reference values in community-dwelling older adults. ESC Heart Fail. 2022;9(3):1703–1712. doi:10.1002/ehf2.13834

23. Zeidan RS, McElroy T, Rathor L, Martenson MS, Lin Y, Mankowski RT. Sex differences in frailty among older adults. Exp Gerontol. 2023;184:112333. doi:10.1016/j.exger.2023.112333

24. Xu L, Zhang J, Shen S, et al. Association Between Body Composition and Frailty in Elder Inpatients. Clin Interv Aging. 2020;Volume 15:313–320. doi:10.2147/CIA.S243211

25. Chao CT, Kovesdy CP, Merchant RA. Sarcopenia, sarcopenic obesity, and frailty in individuals with chronic kidney disease: a comprehensive review. Kidney Res Clin Pract. Published online January 21, 2025. doi:10.23876/j.krcp.24.207

26. Yuan JH, Rifkin DE, Ginsberg C, et al. Difference between kidney function by cystatin C versus creatinine and association with muscle mass and frailty. J Am Geriatr Soc. 2024;72(10):3163–3170. doi:10.1111/jgs.19014

27. Guerville F, De Souto Barreto P, Taton B, et al. Estimated Glomerular Filtration Rate Decline and Incident Frailty in Older Adults. Clin J Am Soc Nephrol. 2019;14(11):1597–1604. doi:10.2215/CJN.03750319

28. Ballew SH, Chen Y, Daya NR, et al. Frailty, Kidney Function, and Polypharmacy: The Atherosclerosis Risk in Communities (ARIC) Study. Am J Kidney Dis. 2017;69(2):228–236. doi:10.1053/j.ajkd.2016.08.034

29. Iacomelli I, Giordano A, Rivasi G, et al. Low Creatinine Potentially Overestimates Glomerular Filtration Rate in Older Fracture Patients: A Plea for an Extensive Use of Cystatin C? Eur J Intern Med. 2021;84:74–79. doi:10.1016/j.ejim.2020.06.016

30. Iversen E, Bodilsen AC, Klausen HH, et al. Kidney function estimates using cystatin C versus creatinine: Impact on medication prescribing in acutely hospitalized elderly patients. Basic Clin Pharmacol Toxicol. 2019;124(4):466–478. doi:10.1111/bcpt.13156

31. Peruzzo S, Ottaviani S, Tagliafico L, et al. Renal function assessment in older people: comparative analysis of estimation equation with serum creatinine. Front Med. 2024;11:1477500. doi:10.3389/fmed.2024.1477500

32. Potok OA, Katz R, Bansal N, et al. The Difference Between Cystatin C– and Creatinine-Based Estimated GFR and Incident Frailty: An Analysis of the Cardiovascular Health Study (CHS). Am J Kidney Dis. 2020;76(6):896–898. doi:10.1053/j.ajkd.2020.05.018

33. Ferguson TW, Komenda P, Tangri N. Cystatin C as a biomarker for estimating glomerular filtration rate: Curr Opin Nephrol Hypertens. 2015;24(3):295–300. doi:10.1097/MNH.0000000000000115

34. Ebert N, Pottel H, Van Der Giet M, Kuhlmann MK, Delanaye P, Schaeffner E. The impact of the new CKD-EPI equation on GFR estimation in the elderly. Dtsch Ärztebl Int. Published online October 14, 2022. doi:10.3238/arztebl.m2022.0258

35. Cavagna I, Fiuzat M, Lala A, et al. Inertia Is Not an Option: Laying the Foundation for a Consensus on the Assessment of Kidney Function in Acute Decompensated Heart Failure. J Card Fail. Published online February 2025:S1071916425000934. doi:10.1016/j.cardfail.2025.01.025

36. Levin A, Ahmed SB, Carrero JJ, et al. Executive summary of the KDIGO 2024 Clinical Practice Guideline for the Evaluation and Management of Chronic Kidney Disease: known knowns and known unknowns. Kidney Int. 2024;105(4):684–701. doi:10.1016/j.kint.2023.10.016

37. Inker LA, Titan S. Measurement and Estimation of GFR for Use in Clinical Practice: Core Curriculum 2021. Am J Kidney Dis. 2021;78(5):736–749. doi:10.1053/j.ajkd.2021.04.016

38. Stevens PE, Ahmed SB, Carrero JJ, et al. KDIGO 2024 Clinical Practice Guideline for the Evaluation and Management of Chronic Kidney Disease. Kidney Int. 2024;105(4):S117-S314. doi:10.1016/j.kint.2023.10.018

39. Brück K, Stel VS, Gambaro G, et al. CKD Prevalence Varies across the European General Population. J Am Soc Nephrol. 2016;27(7):2135–2147. doi:10.1681/ASN.2015050542

40. Bongetti EK, Wilkinson A, Wetmore JB, et al. Association between Urine Albumin and Estimated Glomerular Filtration Rate with Incident Frailty in Healthy Older Adults: Secondary Analysis of the ASPREE Trial Cohort: TH-PO951. J Am Soc Nephrol. 2024;35(10S). doi:10.1681/ASN.202444jyac6d

41. Liu M, He P, Zhou C, et al. Association of urinary albumin:creatinine ratio with incident frailty in older populations. Clin Kidney J. 2022;15(6):1093–1099. doi:10.1093/ckj/sfac002

42. Damanti S, Citterio L, Zagato L, et al. Sarcopenic obesity and pre-sarcopenia contribute to frailty in community-dwelling Italian older people: data from the FRASNET study. BMC Geriatr. 2024;24(1):638. doi:10.1186/s12877-024-05216-6

43. Amiri S, Behnezhad S, Hasani J. Body Mass Index and risk of frailty in older adults: A systematic review and meta-analysis. Obes Med. 2020;18:100196. doi:10.1016/j.obmed.2020.100196

44. Okamura M, Konishi M, Butler J, Kalantar-Zadeh K, Von Haehling S, Anker SD. Kidney function in cachexia and sarcopenia: Facts and numbers. J Cachexia Sarcopenia Muscle. 2023;14(4):1589–1595. doi:10.1002/jcsm.13260

45. Ebert N. Neue Formeln zur Schätzung der Nierenfunktion – Bedeutung für die Dosierung nierengängiger Arzneimittel. Inn Med. 2024;65(3):280–285. doi:10.1007/s00108-023-01649-0

