## Supplementary for "Kidney Function Estimation and its Relationship with Frailty in Older Adults: Insights from SHIP-TREND": Supplementary_Komleva_final.docx

**Supplementary material (Methods): Kidney function assessment,** C-reactive protein, Socio-demographic variables and body composition, Handgrip strength

**Supplementary tables**

**Table 1.** Body composition parameters across frailty phenotypes

**Table 2.** Correlation between frailty score and anthropometric measures

**Table 3.** Linear regression models assessing the association between frailty score, eGFR, muscle mass, and their interaction

**Table 4.** Participant characteristics stratified by frailty and CKD stage

**Table 5.** Prevalence of CKD categories based on eGFR and UACR

**Figure 1.** Frailty prevalence across eGFR categories based on different estimation equations

**Table 6**. Multivariable associations between kidney function and frailty

**Table 6 a**. Association between eGFR MDRD_SCr_ and frailty (frailty score ≥3) adjusted for selected confounders

**Table 6 b.** Associations between measures of kidney function eGFR CKD-EPI_SCr_ and frailty score ≥3 and other confounders: models with UACR

**Table 6 с**. Associations between measures of kidney function eGFR CKD-EPI_SCr_ and frailty score ≥3 and other confounders: models without UACR

**Table 6 d**. Associations between measures of kidney function eGFR CKD-EPI _SCr_ and frailty score ≥3 and other confounders: models with UACR

**Table 6 e**. Associations between measures of kidney function eGFR CKD-EPI_CysC_ and frailty score ≥3 and other confounders: models without UACR

**Table 6 f.** Associations between measures of kidney function eGFR CKD-EPI_CysC_ and frailty score ≥3 and other confounders: models with UACR

**Table 6 g.** Associations between measures of kidney function eGFR EKFC_SCr_ and frailty score ≥3 and other confounders: models without UACR

**Table 6 h.** Associations between measures of kidney function eGFR EKFC_SCr_ and frailty score ≥3 and other confounders: models with UACR

**Table 6 i.** Associations between measures of kidney function eGFR EKFC_CysC_ and frailty score ≥3 and other confounders: models without UACR

**Table 6 j.** Associations between measures of kidney function eGFR EKFC_CysC_ and frailty score ≥3 and other confounders: models with UACR

**Supplementary material (Methods)**

**Kidney function assessment**

Creatinine and Cystatin C were measured in blood serum. In SHIP-Trend, serum creatinine concentrations were determined with a modified kinetic Jaffé method with measurement range 0.14–20.2 mg/dl (Dimension VISTA, Siemens Healthcare Diagnostics, Eschborn, Germany). Serum cystatin C concentrations were measured using a nephelometric assay (Dimension VISTA, Siemens Healthcare Diagnostics, Eschborn, Germany) with detection limit 0.05 mg/l. eGFR was calculated by MDRD_SCr_, CKD-EPI_SCr_, CKD-EPI_CysC_, EKFC_SCr_, EKFC_CysC_ Equation.

The four-variable Modification of Diet in Renal Disease (MDRD) study equation (MDRD) was calculated as : eGFR(crea) = 186.3 × serum creatinine^-1.154^ × age^-0.203^ × (0.742 if female) ^12,13^

The CKD-EPI serum creatinine equation (CKD-EPI_SCr_) was calculated as: 141 × min(Scr/*κ*, 1)*α* × max(Scr/*κ*, 1)−1.209 × 0.993Age [× 1.018 if female] [× 1.159 if black], where Scr is serum creatinine, *κ* is 0.7 for females and 0.9 for males, *α* is −0.329 for females and −0.411 for males, min is the minimum of Scr/*κ* or 1, and max is the maximum of Scr/*κ* or 1. ^15^ Furthermore, cystatin C-based eGFR was calculated using the CKD-EPI cystatin C equation: eGFR(CKD-EPI_CysC_) = 133 × min(serum cystatin C / 0.8, 1)^-0.499^ × max(serum cystatin C / 0.8, 1)^-1.328^ × 0.996^age^ [× 0.932 if female].

The general form of the EKFC eGFR equation is EKFC − eGFR = 107.3/[Biomarker/Q]^α^ × [0.990^(Age−40)^ if age >40 years], with α=0.322 when biomarker/Q is less than 1 and α=1.132 when biomarker/Q is 1 or more^10^.

Urine samples were collected between 7.00 a.m. and 6.00 p.m. The urinary creatinine concentration was measured with a photometric reaction in SHIP-Trend (Dimension VISTA, Siemens Healthcare Diagnostics, Eschborn, Germany). The urinary albumin concentration was measured with nephelometric assays (SHIP-Trend: Dimension VISTA, Siemens Healthcare Diagnostics, Eschborn, Germany). The UACR was calculated using the following equation: UACR (mg/g) = urinary albumin concentration (mg/L)/urinary creatinine concentration (mmol/L)^12,45^.

UACR was stratified into <30 mg/g, 30–299 mg/g, and ≥300 mg/g. CKD was defined as eGFR <60 mL/min/1.73 m2. eGFR was further grouped into: <30, 30–44, 45–59, and ≥60  mL/min/1.73m2.

**C-reactive protein**

In SHIP-Trend, blood samples were taken from the cubital vein of participants in the supine position. Immunonephelometric assays on a Dimension VISTA (Siemens Healthcare Diagnostics, Eschborn, Germany) were used to measure serum hs‐CRP (measuring range 0.016–0.95 mg/dL, CV 3.6–3.8%) ^22^.

**Socio-demographic variables and body composition**

Socio-demographic characteristics were assessed by computer- assisted personal interviews. Height and weight were measured to calculate the body mass index (BMI = weight (kg)/height2 (m2)). Using a multifrequency Nutriguard M device (Data Input, Pöcking, Germany) and the NUTRI4 software (Data Input, Pöcking, Germany) fat free mass was measured in participants without pacemakers. The electrodes were placed on one foot, ankle and hand. Test frequencies were 5, 50 and 100 kHz following the manufacturers instruction. Waist circumference was measured to the nearest 0.1 cm using an inelastic tape midway between the lower rib margin and the iliac crest in the horizontal plane with the subject standing comfortably with weight evenly distributed on both feet ^19,46,47^.

**Handgrip strength**

Handgrip strength was measured by Smedley's Dynamometer, Scandidact, Odder, Denmark. We used the original cut-off values as suggested by Fried et al. ^16^. Standing participants were instructed to keep the upper arm close to the trunk and bring the elbow in a 90° flexion. Afterward they gripped the handle with the maximum effort for 3 seconds, one time with each hand. For further analysis the highest value, whether from the right or left hand, was taken ^19^.

**Supplementary tables**

**Table 1. Body composition parameters across frailty phenotypes**

|  |  | | | **P-value** |
| --- | --- | --- | --- | --- |
|  | **Robust (Frailty-Score = 0)** | **Prefrail (Frailty -score 1-2)** | **Frail (Frailty Score 3-5)** |  |
| Body mass index (kg/m2) | 28.62 [25.98; 31.44] | 29.65 [26.85; 33.05] | 30.78 [28.68; 35.55] | **<0.001** |
| Muscle mass index (kg/m2) | 19.95 [18.03; 22.10] | 20.23 [18.21; 22.37] | 19.95 [18.65; 21.72] | 0.501 |
| Body weight (kg) | 80.15 [71.50; 91.70] | 81.50 [72.30; 93.30] | 78.90 [70.70; 89.90] | 0.129 |
| Muscle weight (kg) | 55.90 [47.10; 65.60] | 54.50 [46.50; 65.00] | 49.30 [44.80; 56.30] | **0.026** |
| Muscle weight / Body weight | 0.71 [0.64; 0.76] | 0.69 [0.62; 0.74] | 0.64 [0.59; 0.73] | **<0.001** |

**Post hoc comparisons of body composition between frailty groups**

|  | **Post hoc analysis: P-value (adjusted)** | | |
| --- | --- | --- | --- |
|  | Robust 0 vs Prefrail 1-2 | Robust 0 vs Frail 3-5 | Prefrail 1-2 vs Frail 3-5 |
| Body mass index (kg/m2) | **<0.001** | 0.150 | 0.633 |
| Muscle weight (kg) | 1.000 | **0.024** | 0.052 |
| Muscle Weight / Body Weight | **0.003** | **0.008** | 0.129 |

**Notes and abbreviations**: Data are presented as median and interquartile range [Q1; Q3], Comparison of BMI, muscle and fat mass, and body composition ratios between robust, pre-frail, and frail participants. Frailty was associated with lower muscle mass. BMI - Body mass index. Adjusted p-values for pairwise comparisons between robust, pre-frail, and frail groups for indicators of body composition, including body mass index, muscle mass, and their relative contributions to body weight. Bold indicates reliable statistical differences (p<0.05).

**Table 2. Correlation between frailty score and anthropometric measures**

|  | **Frailty (score)** | |
| --- | --- | --- |
| Body mass index (kg/m2) | **Pearson r = 0.140** | **P-value <0.001** |
| Muscle mass index (kg/m2) | Pearson r = 0.030 | P-value = 0.257 |
| Muscle weight (kg) | **Pearson r = -0.073** | **P-value = 0.007** |
| Muscle Weight / Body Weight | **Pearson r = -0.127** | **P-value <0.001** |

**Notes:** Pearson correlation coefficients between frailty scores and body composition metrics. Significant positive correlations were observed for fat-related indices, while muscle mass and muscle-to-body weight ratio were inversely associated with frailty. Bold indicates reliable statistical differences (p<0.05).

**Table 3. Linear regression models assessing the association between frailty score, eGFR, muscle mass, and their interaction**

| **eGFR Formula** | **β (eGFR)**  **[95 % CI]** | **eGFR (*p*)** | **β (Muscle mass)**  **[95 % CI]** | **Muscle mass (*p*)** | | **β (Interaction eGFR × muscle mass)**  **[95 % CI]** | **Interaction (eGFR × muscle mass) (*p*)** | | **Model**  **R^2^** |
| --- | --- | --- | --- | --- | --- | --- | --- | --- | --- |
| MDRD _SCr_ | -0.011  [–0.021, –0.001] | **0.037** | -0.014  [-0.028, 0.000] | 0.054 | 0.000  [0.000, 0.000] | | 0.182 | 0.017 | |
| CKD-EPI _SCr_ | -0.015  [–0.027, –0.003] | **0.014** | -0.018  [-0.035, 0.002] | **0.032** | 0.000  [0.000,0.000] | | 0.099 | 0.020 | |
| EKFC _SCr_ | -0.019  [–0.032, –0.005] | **0.008** | -0.019  [-0.036, 0.003] | **0.023** | 0.000  [0.000, 0.000] | | 0.075 | 0.022 | |
| CKD-EPI _Cys C_ | -0.017  [–0.028, –0.006] | **0.002** | -0.019  [-0.036, 0.001] | **0.037** | 0.000  [0.000, 0.000] | | 0.095 | 0.045 | |
| EKFC _Cys C_ | -0.030  [–0.047, –0.013] | **<0.001** | -0.030  [-0.054,-0.006] | **0.016** | 0.000  [0.000, 0.001] | | **0.046** | 0.047 | |

**Notes and abbreviations**: The table presents regression coefficients (β), 95% confidence intervals (CI), and p-values for the associations of eGFR, muscle mass, and their interaction (eGFR × muscle mass) with frailty score. R² represents the proportion of variance in frailty score explained by each model. eGFR: estimated glomerular filtration rate; SCr: serum creatinine; CysC: cystatin C; MDRD: Modification of Diet in Renal Disease; CKD-EPI: Chronic Kidney Disease Epidemiology Collaboration; EKFC: European Kidney Function Consortium.

**Table 4. Participant characteristics stratified by frailty and CKD stage**

| **Variable** | | | **Robust** | **Pre-frail** | **Frail** |
| --- | --- | --- | --- | --- | --- |
| All participants | | | 829 (57.1%) | 591 (40.7%) | 33 (2.3%) |
| **Sex** | | male | 431 (58.4%) | 299 (40.5%) | 8 (1.1%) |
|  |  | female | 398 (55.7%) | 292 (40.8%) | 25 (3.5%) |
| **eGFR: level,**  **n (%)** | **MDRD_SCr_** | <30 | 2 (25%) | 4 (50%) | 2 (25%) |
|  |  | 30–44 | 22 (44.9%) | 26 (53.1%) | 1 (2.0%) |
|  |  | 45–59 | 114 (53.0%) | 95 (44.2%) | 6 (2.8%) |
|  |  | ≥60 | 691 (58.5%) | 466 (39.5%) | 24 (2.0%) |
|  | **CKD-EPI_SCr_** | <30 | 2 (25%) | 4 (50%) | 2 (25%) |
|  |  | 30–44 | 19 (41.3%) | 26 (56.5%) | 1 (2.2%) |
|  |  | 45–59 | 98 (54.4%) | 76 (42.2%) | 6 (3.3%) |
|  |  | ≥60 | 710 (58.2%) | 485 (39.8%) | 24 (2.0%) |
|  | **CKD-EPI_CysC_** | <30 | 1 (20%) | 2 (40%) | 2 (40%) |
|  |  | 30–44 | 6 (23.1%) | 20 (76.9%) | 0 (0.0%) |
|  |  | 45–59 | 35 (44.9%) | 40 (51.3%) | 3 (3.8%) |
|  |  | ≥60 | 787 (58.6%) | 529 (39.4%) | 28 (2.1%) |
|  | **EKFC_SCr_** | <30 | 3 (27.3%) | 6 (54.5%) | 2 (18.2%) |
|  |  | 30–44 | 47 (48.5%) | 48 (49.5%) | 2 (2.1%) |
|  |  | 45–59 | 189 (53.7%) | 154 (43.8%) | 9 (2.6%) |
|  |  | ≥60 | 590 (59.4%) | 383 (38.6%) | 20 (2.0%) |
|  | **EKFC_CysC_** | <30 | 0 (0.0%) | 2 (66.7%) | 1 (33.3%) |
|  |  | 30–44 | 4 (21.1%) | 14 (73.7%) | 1 (5.3%) |
|  |  | 45–59 | 30 (44.1%) | 36 (52.9%) | 2 (2.9%) |
|  |  | ≥60 | 795 (58.3%) | 539 (39.5%) | 29 (2.1%) |
| **UACR level** | | <30 | 495 (56.5%) | 360 (41.1%) | 21 (2.4%) |
|  |  | 30–299 | 134 (57.0%) | 95 (40.4%) | 6 (2.6%) |
|  |  | ≥300 | 17 (47.2%) | 17 (47.2%) | 2 (5.6%) |

**Notes and abbreviations**: Baseline demographic and clinical characteristics of participants categorized as robust, pre-frail, or frail. Data are shown as absolute values and percentages. eGFR stages are presented using five estimation equations (MDRD_SCr_, CKD-EPI_SCr_, CKD-EPI_CysC_, EKFC_SCr_, and EKFC_CysC_). UACR categories reflect albuminuria levels (<30, 30–299, ≥300 mg/g). eGFR: estimated glomerular filtration rate; SCr: serum creatinine; CysC: cystatin C; MDRD: Modification of Diet in Renal Disease; CKD-EPI: Chronic Kidney Disease Epidemiology Collaboration; EKFC: European Kidney Function Consortium. UACR: urine albumin-to-creatinine ratio (mg/g).

**Table 5. Prevalence of CKD categories based on eGFR and UACR**

| **MDRD_SCr_** | | **UACR: mg/g** | | |
| --- | --- | --- | --- | --- |
|  |  | **А1 (<30)** | **A2 (30-299)** | **A3 (≥300)** |
| eGFR:  ml/min/1.73 m^2^ | G1-G2 (≥60) | 724 (63.1%*) | 173 (15.1%) | 19 (1.7%) |
|  | G3a (45–59) | 126 (11.0%) | 40 (3.5%) | 11 (1.0%) |
|  | G3b (30–44) | 24 (2.1%) | 18 (1.6%) | 4 (0.3%) |
|  | G4-G5 (<30) | 2 (0.2%) | 4 (0.3%) | 2 (0.2%) |
| **CKD-EPI_SCr_** | | **UACR: mg/g** | | |
|  |  | **А1 (<30)** | **A2 (30-299)** | **A3 (≥300)** |
| eGFR:  ml/min/1.73 m^2^ | G1-G2 (≥60) | 744 (64.9%*) | 177 (15.4%) | 21 (1.8%) |
|  | G3a (45–59) | 108 (9.4%) | 35 (3.1%) | 11 (1.0%) |
|  | G3b (30–44) | 22 (1.9%) | 19 (1.7%) | 2 (0.2%) |
|  | G4-G5 (<30) | 2 (0.2%) | 4 (0.3%) | 2 (0.2%) |
| **CKD-EPI_CysC_** | | **UACR: mg/g** | | |
|  |  | **А1 (<30)** | **A2 (30-299)** | **A3 (≥300)** |
| **eGFR:**  **ml/min/1.73 m^2^** | **G1-G2 (≥60)** | 823 (71.8%*) | 201 (17.5%) | 25 (2.2%) |
|  | **G3a (45**–**59)** | 41 (3.6%) | 21 (1.8%) | 8 (0.7%) |
|  | **G3b (30**–**44)** | 11 (1.0%) | 11 (1.0%) | 1 (0.1%) |
|  | **G4-G5 (<30)** | 1 (0.1%) | 2 (0.2%) | 2 (0.2%) |
| **EKFC_SCr_** | | **UACR: mg/g** | | |
|  |  | **А1 (<30)** | **A2 (30-299)** | **A3 (≥300)** |
| **eGFR:**  **ml/min/1.73 m^2^** | **G1-G2 (≥60)** | 603 (52.6%*) | 144 (12.6%) | 15 (1.3%) |
|  | **G3a (45**–**59)** | 220 (19.2%) | 55 (4.8%) | 12 (1.0%) |
|  | **G3b (30**–**44)** | 50 (4.4%) | 31 (2.7%) | 7 (0.6%) |
|  | **G4-G5 (<30)** | 3 (0.3%) | 5 (0.4%) | 2 (0.2%) |
| **EKFC_CysC_** | | **UACR: mg/g** | | |
|  |  | **А1 (<30)** | **A2 (30-299)** | **A3 (≥300)** |
| **eGFR:**  **ml/min/1.73 m^2^** | **G1-G2 (≥60)** | 834 (72.7%) | 204 (17.8%) | 28 (2.4%) |
|  | **G3a (45**–**59)** | 32 (2.8%) | 23 (2.0%) | 6 (0.5%) |
|  | **G3b (30**–**44)** | 10 (0.9%) | 7 (0.6%) | 0 (0.0%) |
|  | **G4-G5 (<30)** | 0 (0.0%) | 1 (0.1%) | 2 (0.2%) |

The table shows the number and percentage of participants categorized by eGFR (glomerular filtration rate) and UACR (urinary albumin-to-creatinine ratio) based on five different equations: MDRD_SCr_, CKD-EPI_SCr_, CKD-EPI_CysC_, EKFC_SCr_, and EKFC_CysC_. Results are stratified by albuminuria categories A1 (<30 mg/g), A2 (30–299 mg/g), and A3 (≥300 mg/g), and by eGFR stages: G1-G2 (≥60), G3a (45–59), G3b (30–44), and G4-G5 (<30) mL/min/1.73 m². The analysis was conducted among 1147 participants with assessed frailty, eGFR and UACR. * Percentage of all participants. eGFR: estimated glomerular filtration rate; SCr: serum creatinine; CysC: cystatin C; MDRD: Modification of Diet in Renal Disease; CKD-EPI: Chronic Kidney Disease Epidemiology Collaboration; EKFC: European Kidney Function Consortium. *UACR: urine albumin-to-creatinine ratio (mg/g).*

**Figure 1. Frailty prevalence across eGFR categories based on different estimation equations**

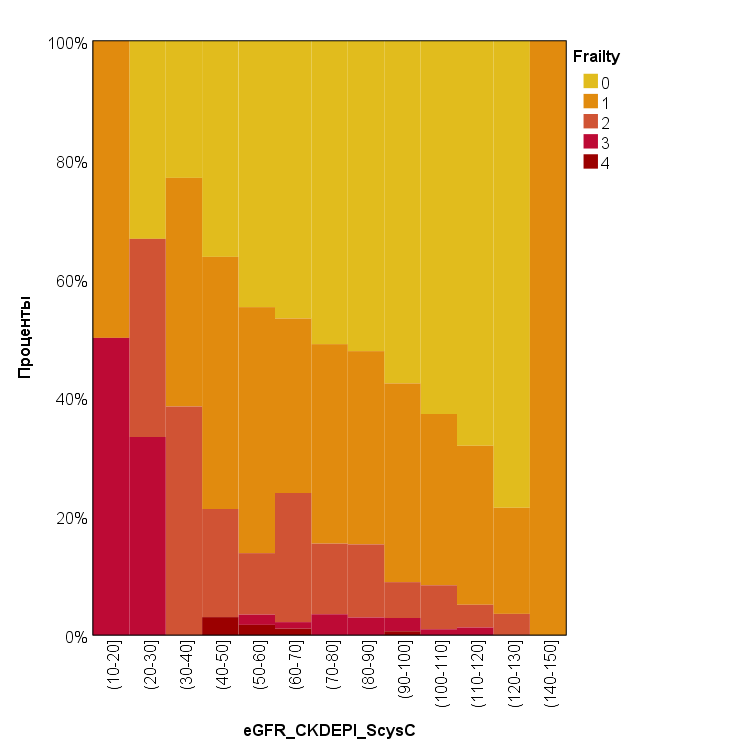

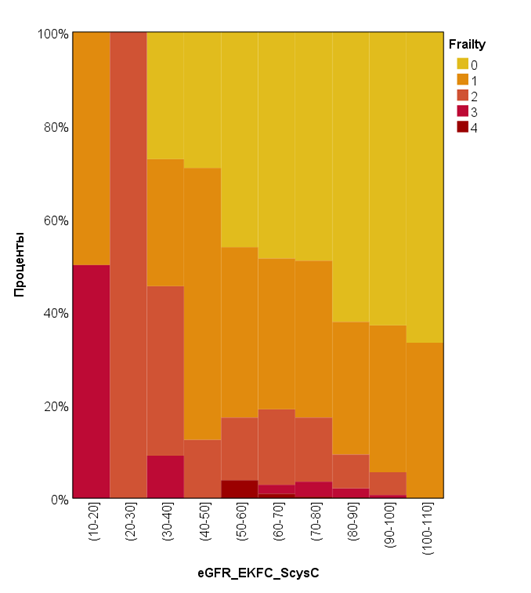

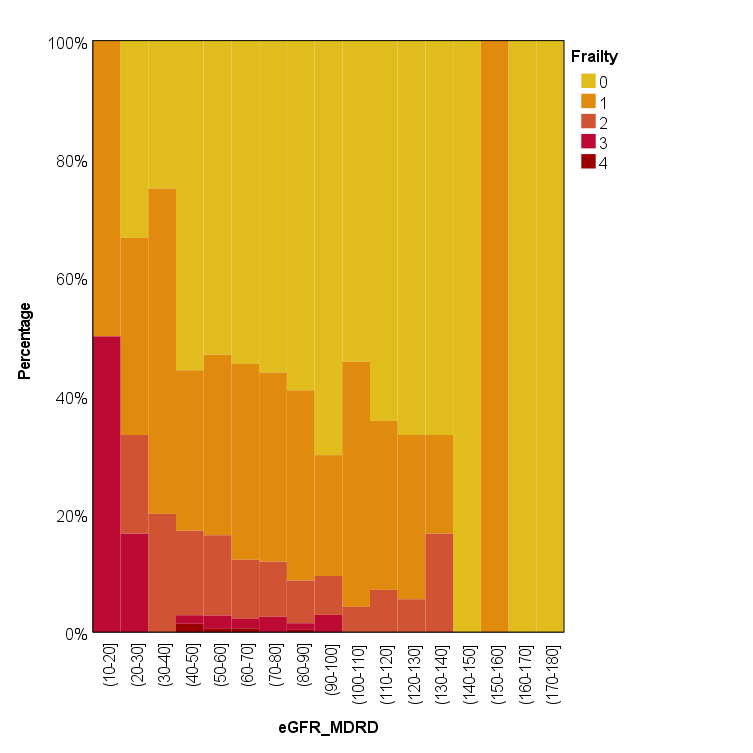

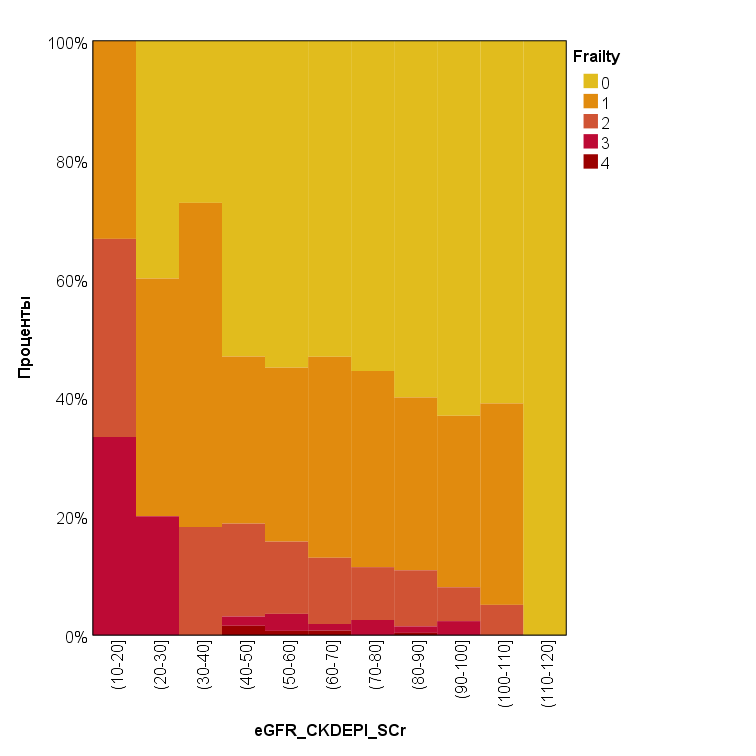

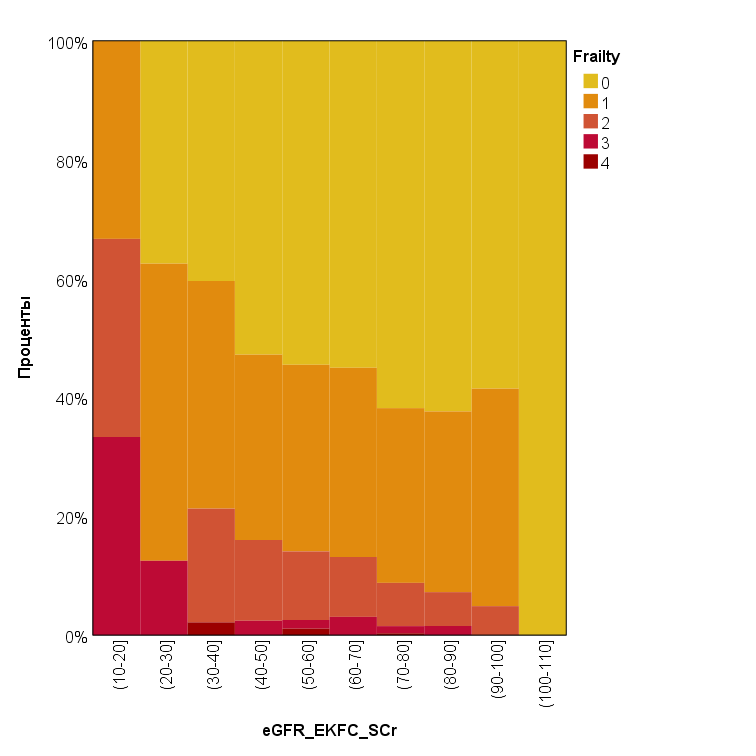

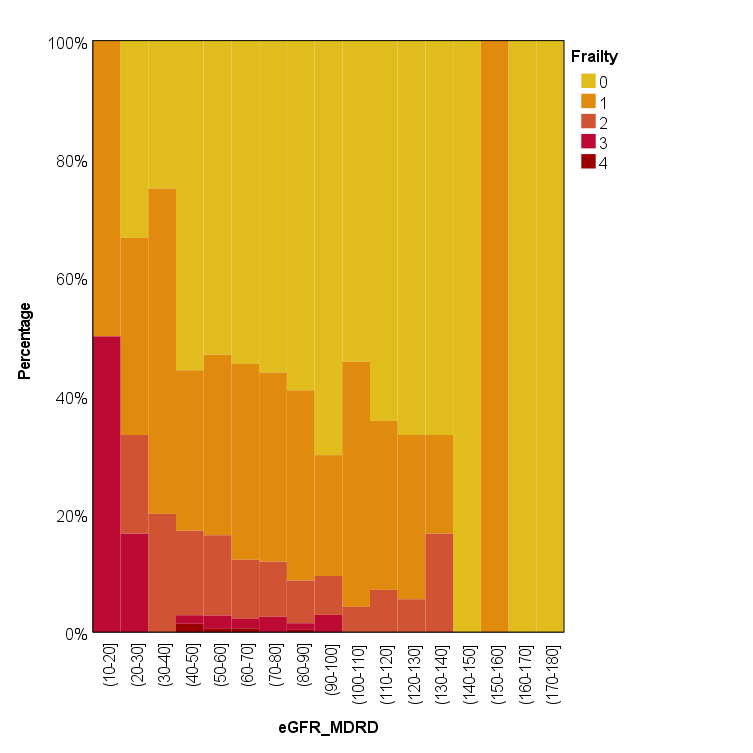

**CKD-EPI_CysC_**

**MDRD_SCr_**

**EKFC_SCr_**

Frailty, %

eGFR, mL/min/1.73m^2^

**CKD-EPI_SCr_**

**EKFC_CysC_**

The figure illustrates the proportion of frail individuals across various eGFR categories (G1–G5) as estimated by five equations: MDRD _SCr_, CKD-EPI _SCr_, CKD-EPI _SCysC_, EKFC _SCr,_ and EKFC _SCysC_. A consistent trend of higher frailty prevalence with declining eGFR was observed across all methods, with the CKD-EPI _SCysC_ and EKFC _SCysC_ equations identifying the highest proportions. eGFR: estimated glomerular filtration rate; SCr: serum creatinine; CysC: cystatin C; MDRD: Modification of Diet in Renal Disease; CKD-EPI: Chronic Kidney Disease Epidemiology Collaboration; EKFC: European Kidney Function Consortium.

**Table 6**. **Multivariable associations between kidney function and frailty**

Table 6 a. Association between eGFR MDRD_SCr_ and frailty (frailty score ≥3) adjusted for selected confounders

| **Confounders included** | | | **sex, age** | **comorbidity** | **sex, age, comorbidity** |
| --- | --- | --- | --- | --- | --- |
| **Adjusted OR [95% CI],**  **p-value** | eGFR;  mL/min/1.73m^2^ | <30 (vs ≥60*) | **8.82 [1.59;49,00],**  **P=0.013** | **9.38 [1.64;53.42],**  **P=0.012** | **6,68 [1,18;37,90],**  **P = 0.032** |
|  |  | 30-44 (vs ≥60) | 0.67 [0.09;5.18],  P=0.699 | 0.67 [0.09;5.25],  P=0.707 | 0,50 [0,06;4,06],  P = 0.520 |
|  |  | 45-59 (vs ≥60) | 0.81 [0.31;2.08],  P=0.668 | 1.11 [0.44;2.81],  P=0.824 | 0,73 [0,28;1,88],  P = 0.513 |
|  | Sex (female vs male) | | **3.68 [1.62; 8.33]; p = 0.002** | **-** | **3.45 [1.52;7.84],**  **P = 0.003** |
|  | Age (years) | | **1.1 [1.04; 1.17];**  **p = 0.002** | **-** | **1.09 [1.03;1.16],**  **P = 0.005** |
|  | Comorbidity (number of chronic diseases) | | - | **1.36 [1.08;1.72],**  **P=0.010** | 1.26 [0.99;1.61],  P=0.055 |
| **Model characteristics** | | | **P (model) <0.001**  Pseudo R2 = 0.083 | **P (model) = 0.009**  Pseudo R2 = 0.043 | **P (model) <0.001**  Pseudo R2 = 0.095 |

Table 6 b. Associations between measures of kidney function eGFR CKD-EPI_SCr_ and frailty score ≥3 and other confounders: models with UACR

| **Multivariable logistic regression** | | | **eGFR &**  **UACR, sex, age** | **eGFR**  **& UACR, comorbidity** | **eGFR &**  **UACR, sex, age, comorbidity** |
| --- | --- | --- | --- | --- | --- |
| **Adjusted OR [95% CI],**  **P-value** | eGFR;  mL/min/1.73m^2^ | <30 (vs ≥60*) | **8.21 [1.38;48.79],**  **p = 0.021** | **9.29 [1.52;56.75],**  **p = 0.016** | **6.73 [1.12; 40.54]**  **p = 0.038** |
|  |  | 30-44 (vs ≥60) | 0.64 [0.08;5.12],  p=0.675 | 0.72 [0.09;5.70],  p = 0.753 | 0.51 [0.06; 4.24]  p = 0.534 |
|  |  | 45-59 (vs ≥60) | 0.92 [0.35;2.42],  p =0.861 | 1.28 [0.49;3.33],  P = 0.617 | 0.82 [0.31; 2.19]  p = 0.692 |
|  | UACR (≥300 mg/g vs <300 mg/g*) | | 1.45 [0.29; 7.24]; p = 0.650 | 1.36 [0.27;6.79],  p=0.709 | 1.38 [0.28;6.86],  P = 0.694 |
|  | Sex (female vs male) | | **4.15 [1.73; 9.95]; p = 0.001** | - | **3.96 [1.65;9.54],**  **P = 0.002** |
|  | Age (years) | | **1.10 [1.03; 1.17]; p = 0.007** | - | **1.09 [1.02;1.16],**  **P = 0.013** |
|  | Comorbidity (number of chronic diseases) | | - | 1.28 [0.99;1.65],  p=0.055 | 1.20 [0.93;1.56],  P = 0.158 |
| **Model characteristics** | | | **P (model) <0.001**  Pseudo R2 = 0.092 | **P (model) = 0.096**  Pseudo R2 = 0.040 | **P (model) <0.001**  Pseudo R2 = 0.099 |

Table 6 с. Associations between measures of kidney function eGFR CKD-EPI_SCr_ and frailty score ≥3 and other confounders: models without UACR

| **Multivariable logistic regression** | | | **eGFR &**  **sex, age** | **eGFR &**  **comorbidity** | **eGFR &**  **sex, age, comorbidity** |
| --- | --- | --- | --- | --- | --- |
| **Adjusted OR [95% CI],**  **P-value** | eGFR;  mL/min/1.73m^2^ | <30 (vs ≥60*) | **9.260 [1.668; 51.412]; p = 0.011** | **9.909 [1.747; 56.220]; p = 0.010** | **7.099 [1.252; 40.241]; p = 0.027** |
|  |  | 30-44 (vs ≥60) | 0.726 [0.093; 5.686];  p = 0.760 | 0.739 [0.095; 5.761];  p = 0.773 | 0.549 [0.068; 4.458];  p = 0.575 |
|  |  | 45-59 (vs ≥60) | 0.976 [0.376; 2.531];  p = 0.959 | 1.399 [0.553; 3.536];  p = 0.478 | 0.890 [0.341; 2.326];  p = 0.813 |
|  | Sex (female vs male) | | **3.603 [1.591; 8.155]; p = 0.002** | - | **3.366 [1.481; 7.651]; p = 0.004** |
|  | Age (years) | | **1.101 [1.034; 1.171]; p = 0.003** | - | **1.090 [1.024; 1.162]; p = 0.007** |
|  | Comorbidity (number of chronic diseases) | | - | **1.350 [1.069; 1.705]; p = 0.012** | 1.254 [0.988; 1.593]; p = 0.063 |
| **Model characteristics** | | | **P (model) <0.001**  Pseudo R2 = 0.091 | **P (model) = 0.008**  Pseudo R2 = 0.049 | **P (model) <0.001**  Pseudo R2 = 0.103 |

Table 6 d. Associations between measures of kidney function eGFR CKD-EPI_SCr_ and frailty score ≥3 and other confounders: models with UACR

| **Multivariable logistic regression** | | | **eGFR &**  **UACR, sex, age** | **eGFR**  **& UACR, comorbidity** | **eGFR &**  **UACR, sex, age, comorbidity** |
| --- | --- | --- | --- | --- | --- |
| **Adjusted OR [95% CI],**  **P-value** | eGFR;  mL/min/1.73m^2^ | <30 (vs ≥60*) | **7.972 [1.374; 46.243]; p = 0.021** | **9.345 [1.573; 55.532] p = 0.014** | **6.540 [1.109; 38.549] p = 0.038** |
|  |  | 30-44 (vs ≥60) | 0.700 [0.087; 5.611]; p = 0.737 | 0.784 [0.099; 6.197]  p = 0.818 | 0.562 [0.068; 4.680]  p = 0.594 |
|  |  | 45-59 (vs ≥60) | 1.093 [0.412; 2.901]; p = 0.858 | 1.536 [0.590; 3.997]  p = 0.379 | 1.001 [0.373; 2.687]  p = 0.998 |
|  | UACR (≥300 mg/g vs <300 mg/g*) | | 1.625 [0.336; 7.855]; p = 0.546 | 1.453 [0.299; 7.060]  p = 0.643 | 1.578 [0.329; 7.581]  p = 0.569 |
|  | Sex (female vs male) | | **4.035 [1.688; 9.650]; p = 0.002** | - | **3.846 [1.603; 9.227]**  **p = 0.003** |
|  | Age (years) | | **1.089 [1.019; 1.165]; p = 0.012** | - | **1.083 [1.012; 1.159]**  **p = 0.021** |
|  | Comorbidity (number of chronic diseases) | | - | 1.269 [0.986; 1.633]  p = 0.065 | 1.188 [0.919; 1.536]  p = 0.189 |
| **Model characteristics** | | | **P (model) <0.001**  Pseudo R2 = 0.101 | **P (model) = 0.046**  Pseudo R2 = 0.046 | **P (model) <0.001**  Pseudo R2 = 0.108 |

Table 6 e. Associations between measures of kidney function eGFR CKD-EPI_CysC_ and frailty score ≥3 and other confounders: models without UACR

| **Multivariable logistic regression** | | | **eGFR &**  **sex, age** | **eGFR &**  **comorbidity** | **eGFR &**  **sex, age, comorbidity** |
| --- | --- | --- | --- | --- | --- |
| **Adjusted OR [95% CI],**  **P-value** | eGFR;  mL/min/1.73m^2^ | <30 (vs ≥60*) | **24.073 [3.299; 175.654]; p = 0.002** | **17.528 [2.572; 119.425]; p = 0.003** | **17.111 [2.491; 117.535]; p = 0.004** |
|  |  | 30-44 (vs ≥60) | NA**;  p = 0.998 | NA**;  p = 0.998 | NA**;  p = 0.998 |
|  |  | 45-59 (vs ≥60) | 1.045 [0.295; 3.697]; p = 0.946 | 1.426 [0.413; 4.926]; p = 0.575 | 0.895 [0.249; 3.223]; p = 0.866 |
|  | Sex (female vs male) | | **3.800 [1.665; 8.675]; p = 0.002** | - | **3.624 [1.575; 8.338]; p = 0.002** |
|  | Age (years) | | **1.098 [1.033; 1.168]; p = 0.003** | - | **1.087 [1.021; 1.158]; p = 0.009** |
|  | Comorbidity (number of chronic diseases) | | - | **1.352 [1.071; 1.709]; p = 0.011** | 1.257 [0.989; 1.597]; p = 0.061 |
| **Model characteristics** | | | **P (model) <0.001**  Pseudo R2 = 0.105 | **P (model) = 0,002**  Pseudo R2 = 0.059 | **P (model) <0.001**  Pseudo R2 = 0.117 |

Table 6 f. Associations between measures of kidney function eGFR CKD-EPI_CysC_ and frailty score ≥3 and other confounders: models with UACR

| **Multivariable logistic regression** | | | | **eGFR &**  **UACR, sex, age** | **eGFR**  **& UACR, comorbidity** | **eGFR &**  **UACR, sex, age, comorbidity** |
| --- | --- | --- | --- | --- | --- | --- |
| **Adjusted OR [95% CI],**  **P-value** | | eGFR;  mL/min/1.73m^2^ | <30 (vs ≥60*) | **21.197 [2.545; 176.536]; p = 0.005** | **16.452 [2.187; 123.779]; p = 0.007** | **16.355 [2.089; 128.038]; p = 0.008** |
|  |  |  | 30-44 (vs ≥60) | NA**; p = 0.998 | NA**; p = 0.998 | NA**; p = 0.998 |
|  |  |  | 45-59 (vs ≥60) | 1.040 [0.285; 3.792]; p = 0.952 | 1.479 [0.417; 5.247]; p = 0.545 | 0.912 [0.246; 3.384]; p = 0.890 |
|  |  | UACR (≥300 mg/g vs <300 mg/g*) | | 1.279 [0.226; 7.232]; p = 0.780 | 1.243 [0.230; 6.729]; p = 0.801 | 1.261 [0.229; 6.942]; p = 0.790 |
|  |  | Sex (female vs male) | | **4.320 [1.786; 10.448]; p = 0.001** | - | **4.182 [1.718; 10.182]; p = 0.002** |
|  |  | Age (years) | | **1.089 [1.019; 1.163]; p = 0.012** | - | **1.081 [1.011; 1.156]; p = 0.022** |
|  |  | Comorbidity (number of chronic diseases) | | - | 1.274 [0.990; 1.639]; p = 0.060 | 1.195 [0.923; 1.545]; p = 0.176 |
|  | **Model characteristics** | | | **P (model) <0.001**  Pseudo R2 = 0.117 | **P (model) = 0,017**  Pseudo R2 = 0.057 | **P (model) <0.001**  Pseudo R2 = 0.124 |

Table 6 g. Associations between measures of kidney function eGFR EKFC_SCr_ and frailty score ≥3 and other confounders: models without UACR

| **Multivariable logistic regression** | | | **eGFR &**  **sex, age** | **eGFR &**  **comorbidity** | **eGFR &**  **sex, age, comorbidity** |
| --- | --- | --- | --- | --- | --- |
| **Adjusted OR [95% CI],**  **P-value** | eGFR;  mL/min/1.73m^2^ | <30 (vs ≥60*) | 4.618 [0.852; 25.021]; p = 0.076 | **5.848 [1.070; 31.946]; p = 0.041** | 3.174 [0.536; 18.784]; p = 0.203 |
|  |  | 30-44 (vs ≥60) | 0.484 [0.105; 2.233]; p = 0.352 | 0.701 [0.156; 3.145]; p = 0.642 | 0.388 [0.082; 1.833]; p = 0.232 |
|  |  | 45-59 (vs ≥60) | 0.709 [0.304; 1.650]; p = 0.424 | 1.119 [0.500; 2.504]; p = 0.784 | 0.671 [0.286; 1.573]; p = 0.359 |
|  | Sex (female vs male) | | **3.738 [1.654; 8.450]; p = 0.002** | - | **3.445 [1.517; 7.822]; p = 0.003** |
|  | Age (years) | | **1.113 [1.044; 1.186]; p = 0.001** | - | **1.103 [1.034; 1.177]; p = 0.003** |
|  | Comorbidity (number of chronic diseases) | | - | **1.369 [1.084; 1.729]; p = 0.008** | 1.266 [0.996; 1.611]; p = 0.054 |
| **Model characteristics** | | | **P (model) <0.001**  Pseudo R2 = 0.090 | **P (model) = 0,016**  Pseudo R2 = 0.043 | **P (model) <0.001**  Pseudo R2 = 0.103 |

Table 6 h. Associations between measures of kidney function eGFR EKFC_SCr_ and frailty score ≥3 and other confounders: models with UACR

| **Multivariable logistic regression** | | | **eGFR &**  **UACR, sex, age** | **eGFR**  **& UACR, comorbidity** | **eGFR &**  **UACR, sex, age, comorbidity** |
| --- | --- | --- | --- | --- | --- |
| **Adjusted OR [95% CI],**  **P-value** | eGFR;  mL/min/1.73m^2^ | <30 (vs ≥60*) | 4.180 [0.729; 23.978]; p = 0.109 | **5.865 [1.011; 34.005]; p = 0.049** | 3.091 [0.501; 19.071]; p = 0.224 |
|  |  | 30-44 (vs ≥60) | 0.481 [0.101; 2.279]; p = 0.356 | 0.709 [0.154; 3.267]; p = 0.659 | 0.395 [0.081; 1.931]; p = 0.251 |
|  |  | 45-59 (vs ≥60) | 0.693 [0.279; 1.720]; p = 0.429 | 1.113 [0.470; 2.635]; p = 0.807 | 0.659 [0.264; 1.644]; p = 0.371 |
|  | UACR (≥300 mg/g vs <300 mg/g*) | | 1.898 [0.402; 8.956]; p = 0.418 | 1.737 [0.369; 8.182]; p = 0.485 | 1.862 [0.398; 8.710]; p = 0.430 |
|  | Sex (female vs male) | | **4.271 [1.785; 10.222]; p = 0.001** | - | **4.023 [1.674; 9.669]; p = 0.002** |
|  | Age (years) | | **1.102 [1.029; 1.181]; p = 0.005** | - | **1.096 [1.023; 1.175]; p = 0.010** |
|  | Comorbidity (number of chronic diseases) | | - | **1.291 [1.003; 1.660]; p = 0.047** | 1.204 [0.929; 1.559]; p = 0.161 |
| **Model characteristics** | | | **P (model) <0.001**  Pseudo R2 = 0.101 | P (model) = 0,081  Pseudo R2 = 0.040 | **P (model) <0.001**  Pseudo R2 = 0.109 |

Table 6 i. Associations between measures of kidney function eGFR EKFC_CysC_ and frailty score ≥3 and other confounders: models without UACR

| **Multivariable logistic regression** | | | **eGFR &**  **sex, age** | **eGFR &**  **comorbidity** | **eGFR &**  **sex, age, comorbidity** |
| --- | --- | --- | --- | --- | --- |
| **Adjusted OR [95% CI],**  **P-value** | eGFR;  mL/min/1.73m^2^ | <30 (vs ≥60*) | **22.386 [1.752; 286.105]; p = 0.017** | 11.640 [0.948; 142.856]; p = 0.055 | **14.796 [1.186; 184.530]; p = 0.036** |
|  |  | 30-44 (vs ≥60) | 1.741 [0.215; 14.114]; p = 0.604 | 1.705 [0.211; 13.808]; p = 0.617 | 1.393 [0.168; 11.530]; p = 0.759 |
|  |  | 45-59 (vs ≥60) | 0.829 [0.184; 3.726]; p = 0.806 | 1.018 [0.231; 4.478]; p = 0.982 | 0.698 [0.152; 3.202]; p = 0.643 |
|  | Sex (female vs male) | | **3.831 [1.687; 8.697]; p = 0.001** | - | **3.543 [1.549; 8.104]; p = 0.003** |
|  | Age (years) | | **1.103 [1.037; 1.172]; p = 0.002** | - | **1.092 [1.026; 1.162]; p = 0.006** |
|  | Comorbidity (number of chronic diseases) | | - | **1.370 [1.086; 1.728]; p = 0.008** | 1.248 [0.982; 1.585];  p = 0.070 |
| **Model characteristics** | | | **P (model) <0.001**  Pseudo R2 = 0.088 | **P (model) = 0.022**  Pseudo R2 = 0.040 | **P (model) <0.001**  Pseudo R2 = 0.100 |

Table 6 j. Associations between measures of kidney function eGFR EKFC_CysC_ and frailty score ≥3 and other confounders: models with UACR

| **Multivariable logistic regression** | | | **eGFR &**  **UACR, sex, age** | **eGFR**  **& UACR, comorbidity** | **eGFR &**  **UACR, sex, age, comorbidity** |
| --- | --- | --- | --- | --- | --- |
| **Adjusted OR [95% CI],**  **P-value** | eGFR;  mL/min/1.73m^2^ | <30 (vs ≥60*) | **17.991 [1.040; 311.302]; p = 0.047** | 9.376 [0.634; 138.624]; p = 0.103 | 13.243 [0.808; 217.150]; p = 0.070 |
|  |  | 30-44 (vs ≥60) | 1.974 [0.239; 16.277]; p = 0.528 | 1.938 [0.237; 15.846]; p = 0.537 | 1.671 [0.198; 14.072]; p = 0.637 |
|  |  | 45-59 (vs ≥60) | 0.849 [0.185; 3.901]; p = 0.834 | 1.039 [0.231; 4.670]; p = 0.960 | 0.735 [0.156; 3.456]; p = 0.697 |
|  | UACR (≥300 mg/g vs <300 mg/g*) | | 1.314 [0.222; 7.766]; p = 0.763 | 1.483 [0.276; 7.970]; p = 0.646 | 1.307 [0.229; 7.462]; p = 0.764 |
|  | Sex (female vs male) | | **4.360 [1.814; 10.476]; p = 0.001** | - | **4.108 [1.698; 9.940]; p = 0.002** |
|  | Age (years) | | **1.093 [1.024; 1.168]; p = 0.008** | - | **1.086 [1.016; 1.161]; p = 0.016** |
|  | Comorbidity (number of chronic diseases) | | - | **1.294 [1.009; 1.660]; p = 0.042** | 1.183 [0.915; 1.528]; p = 0.199 |
| **Model characteristics** | | | **P (model) = 0.001**  Pseudo R2 = 0.098 | P (model) = 0.123  Pseudo R2 = 0.036 | **P (model) = 0.001**  Pseudo R2 = 0.104 |

**Notes and abbreviations**: Logistic regression models evaluating the association between reduced kidney function (eGFR <60 mL/min/1.73 m²) and frailty (defined as score ≥3). Separate models were constructed for each eGFR estimation equation (MDRD_SCr_, CKD-EPI_SCr_, CKD-EPI_CysC_, EKFC_SCr_, EKFC_CysC_), with and without adjustment for urinary albumin-to-creatinine ratio (UACR). Covariates included sex, age, and number of comorbidities. Significant associations were observed particularly in the lowest eGFR category (<30), most notably when using CysC-based equations. * reference value. Bold indicates reliable statistical differences (p<0.05).

eGFR: estimated glomerular filtration rate; SCr: serum creatinine; CysC: cystatin C; MDRD: Modification of Diet in Renal Disease; CKD-EPI: Chronic Kidney Disease Epidemiology Collaboration; EKFC: European Kidney Function Consortium; UACR: urine albumin-to-creatinine ratio (mg/g).
